# Acute promyelocytic leukemia with torque teno mini virus (TTMV)::*RARA* fusion: an approach to screening and diagnosis

**DOI:** 10.1101/2024.03.29.24304729

**Authors:** Harrison K. Tsai, Mark F. Sabbagh, Meagan Montesion, Erik A. Williams, Arnaldo Arbini, Daniel R. Boué, Emily M. Harris, Franziska Wachter, Leslie Grimmett, Andrew E. Place, Fabienne Lucas, Valentina Nardi, Annette S. Kim, Carlo Brugnara, Barbara Degar, Jessica Pollard, Marian H. Harris, Jacob R. Bledsoe

**Affiliations:** Department of Pathology, Boston Children’s Hospital, Harvard Medical School, Boston, MA, USA; Department of Pathology, Brigham & Women’s Hospital, Harvard Medical School, Boston, MA, USA; Department of Pathology, Massachusetts General Hospital, Harvard Medical School, Boston, MA, USA; Foundation Medicine Inc., Boston, MA, USA; Department of Pathology and Laboratory Medicine, University of Miami, Sylvester Comprehensive Cancer Center, and Jackson Memorial Hospitals, Miami, FL, USA; Department of Pathology, NYU Grossman School of Medicine, New York City, NY, USA; Department of Pathology & Laboratory Medicine, Nationwide Children’s Hospital and The Ohio State University, Columbus, OH, USA; Dana Farber/Boston Children’s Cancer and Blood Disorders Center, Boston, MA, USA; Department of Laboratory Medicine, Boston Children’s Hospital, Harvard Medical School, Boston, MA, USA

## Abstract

Acute promyelocytic leukemia (APL) with variant *RARA* translocation is linked to over 15 partner genes. Recent publications encompassing six cases have expanded the spectrum of *RARA* partners to torque teno mini virus (TTMV). This entity is likely under-recognized due to lack of clinician and pathologist familiarity, inability to detect the fusion using routine testing modalities, and informatic challenges in its recognition within next-generation sequencing (NGS) data. We describe a clinicopathologic approach and provide necessary tools to screen and diagnose APL with TTMV::*RARA* using existing clinical DNA or RNA-based NGS assays, which led to identification of four cases, all without other known cytogenetic/molecular drivers. One was identified prospectively and three retrospectively, including two from custom automated screening of multiple data sets (50,257 cases of hematopoietic malignancy, including 4809 acute myeloid leukemia (AML)/myeloid sarcoma/APL cases). Two cases presented as myeloid sarcoma, including one with multiple relapses after AML-type chemotherapy and hematopoietic stem cell transplant (HSCT). Two cases presented as leukemia, had a poor response to induction chemotherapy, but achieved remission upon re-induction (including all-trans retinoic acid (ATRA) in one case) and subsequent HSCT. Neoplastic cells demonstrated features of APL including frequent azurophilic granules and dim/absent CD34 and HLA-DR expression. *RARA* rearrangement was not detected by karyotype or FISH. Custom analysis of NGS fusion panel data identified TTMV::*RARA* rearrangements, and in the prospectively identified case, facilitated monitoring in sequential bone marrow samples. APL with TTMV::*RARA* is a rare leukemia with a high rate of treatment failure in described cases. The diagnosis should be considered in leukemias with features of APL that lack detectable *RARA* fusions and other drivers, and may be confirmed by appropriate NGS tests with custom informatics. Incorporation of ATRA may have a role in treatment but requires accurate recognition of the fusion for appropriate classification as APL.

## Introduction

Acute promyelocytic leukemia (APL) is a distinct type of childhood and adult leukemia that is characterized by the presence of a *PML::RARA* gene fusion, most commonly in the form of the diagnostic t(15;17)(q24;q21) translocation. APL with variant *RARA* translocation to genes other than *PML* occurs rarely and is acknowledged as a subtype of APL by the WHO classification of hematolymphoid tumors.^1,2^ The diagnosis of APL depends on recognition of characteristic clinical and pathologic features including the tendency to present with coagulopathy, promyelocytic morphology, and a typical flow cytometry immunophenotype that includes lack of HLA-DR and CD34 expression in most cases. These features prompt cytogenetic and molecular testing to confirm the presence of *PML::RARA* or alternate *RARA* fusion. Rapid recognition of APL is necessary to identify patients who will benefit from immediate initiation of all-trans retinoic acid (ATRA) to limit their risk of potentially life-threatening bleeding related to disseminated intravascular coagulation. Accurate diagnosis is critically important as the treatment strategy for APL differs from that of other types of AML and centers on therapy with ATRA and arsenic trioxide (ATO).^3^ Among pediatric patients with APL, the use of ATRA and ATO has been shown to safely allow elimination or substantial reduction of exposure to cytotoxic chemotherapy with non-inferior outcomes.^4^

Torque teno mini virus (TTMV) is a non-enveloped, circular, single-stranded DNA virus in the Anellovirus family.^5^ Anelloviruses are the most abundant eukaryotic viruses in the human virome, and it has been estimated that 90-100% of humans are infected at an early age.^6,7^ Anelloviruses have been detected in almost all human body fluids suggesting that the virus infects many different cell types.^6^ Prior studies have identified particular TTMV types in the serum or plasma of patients with various hematologic malignancies including lymphomas and multiple myeloma, suggesting a possible etiologic link.^8,9^

Recent reports have documented the fascinating occurrence of fusions between *RARA* and TTMV as a cause of APL with clinicopathologic features similar to typical APL, including response to APL-type therapy.^10,11^ To our knowledge only six prior cases have been documented in the peer-reviewed literature,^10,12–15^ with variable clinical and molecular characterization. While the occurrence of APL with TTMV::*RARA* is thought to be uncommon, lack of awareness by clinicians and pathologists, and limitations of current clinical testing modalities may result in under-recognition of this entity.

We report four cases of APL with TTMV::*RARA* fusions that were not detected by routine cytogenetic and molecular testing, but were identified by custom analysis of fusion data from two commercial clinical platforms. We highlight clinicopathologic features useful to recognize these cases and describe an approach to screening and identification. Using an automated method, we screen several large cohorts of acute leukemia and other hematopoietic malignancies for TTMV and Anellovirus integration, confirming the rarity of this entity and allowing initial estimations of its frequency in pediatric and adult populations.

## Materials and Methods

### Case identification and screening of historic cases

The study was conducted in accordance with the Helsinki Declaration and with approval of Institutional Review Boards. The first stage of the study assembled a cohort of leukemias both retrospectively (review of pathology files) and prospectively (during routine clinical practice) with clinicopathologic features suggestive of APL but with negative *PML::RARA* and/or *RARA* break-apart fluorescent in situ hybridization (FISH) testing. These candidates were then tested by a clinical fusion panel supplemented by custom informatic methods designed to screen for TTMV::*RARA*.

Features suggestive of APL included:

a. Promyelocyte-like or leukemic cell morphology with heavy cytoplasmic granulation.
b. APL-like flow cytometry immunophenotype (negative for HLA-DR and negative or partial CD34).
c. Cases in which a diagnosis of APL was initially considered by the pathologist but subsequently excluded by *PML::RARA* and/or *RARA* break-apart FISH.

The second stage of the study screened broad cohorts of hematologic malignancy samples with existing clinical fusion panel data through application of our custom informatics methods.

### NGS panel testing for fusions

RNA-based fusion panels: Total nucleic acid extracted from peripheral blood, bone marrow, or extramedullary disease sites was tested at the Clinical Laboratory Improvement Amendments (CLIA)-certified Laboratory of Molecular Pediatric Pathology at Boston Children’s Hospital (BCH) between 6/11/2018 and 9/29/2023 or at the CLIA-certified Center for Integrated Diagnostics at Massachusetts General Hospital (MGH) between 5/18/2017 and 8/3/2023 using clinically validated targeted RNA next generation sequencing (NGS) panels based on anchored multiplex PCR (AMP) designed principally for fusion detection.^16^ The assays include *RARA* gene specific primers targeting the 5’ ends of exons 1-9 as 3’ fusion partners and the 3’ ends of exons 6-7 as 5’ fusion partners, where exon annotations are relative to the *RARA* transcript NM_000964.4. The BCH assay utilized the commercially available ArcherDx FusionPlex Heme v2 kit (Integrated DNA Technologies, Coralville, Iowa, USA; Product Number AB0063), with library preparation performed according to recommended assay protocols including the use of unique molecular identifiers (UMI). The MGH assay also utilized primers from ArcherDx within an internally developed protocol for library preparation. Samples underwent 2 x 151 bp paired-end Illumina sequencing followed by bioinformatics processing consisting of Archer Analysis v6.2.7 (AA) under default parameters at BCH and an internally developed computational pipeline at MGH. NGS samples not passing clinical quality control metrics were excluded.

DNA-based panel: Comprehensive genomic profiling (CGP) was performed by Foundation Medicine Inc (FMI) between 12/28/2013 and 3/7/2023 using the FoundationOne®Heme assay as part of routine clinical care in a CLIA-certified, College of American Pathologists-accredited, New York State-approved Laboratory, as previously described.^17^ DNA extracted from peripheral blood, bone marrow, or formalin-fixed, paraffin-embedded (FFPE) tissue underwent a hybridization-capture, adapter ligation-based library preparation, followed by 2 x 49 bp paired-end Illumina sequencing to a median exon coverage depth of at least 500x for all exons and select introns of over 400 cancer-related genes including *RARA*. Analysis for genomic alterations, including short variant alterations (base substitutions as well as small insertions and deletions), copy number alterations (amplifications and homozygous deletions), and gene rearrangements, was performed as previously described.^18^ Approval for inclusion of these samples in this study, including a waiver of informed consent and the Health Insurance Portability and Accountability Act (HIPAA) waiver of authorization, was obtained from the Western Institutional Review Board (Protocol No. 20152817).

The cohorts tested by NGS panels for rearrangements consisted of:

1. BCH cohort: n=510 pediatric hematologic samples comprised of 110 AML, 256 B-ALL, 40 T-ALL, 12 lymphoma, 8 MPAL, 9 MDS, 5 CML, 4 MPN, 30 remission, and 36 other/unknown.
2. MGH cohort: n=2174 hematologic samples including 862 adult (age >18 years) AML and 10 pediatric AML.
3. FMI cohort: n=47,573 total cases of hematopoietic malignancies including 3112 adult and 650 pediatric AML/myeloid sarcoma, 53 adult and 12 pediatric APL, and 2065 adult and 1183 pediatric other leukemias, respectively.

### TTMV::*RARA* custom analysis

Manual review for TTMV::*RARA* was performed by visually screening binary alignment map (BAM) files in Integrative Genomics Viewer (IGV) for unusual reads that either (i) aligned partially to *RARA* intron 2 with 5’ soft clipping not found in a selection of other samples with established non-APL diagnoses, or (ii) were spliced to *RARA* exon 3 but with 5’ soft clipping patterns not seen in the selected other samples. The 5’ soft clips were then further evaluated using the web-based interface for nucleotide blast to the nr/nt nucleotide database (https://blast.ncbi.nlm.nih.gov; last accessed 9/28/2023). This approach was effective for evaluating a small number of samples, as long as the underlying BAM files were generated by a modern local aligner to the human genome and employed relatively relaxed alignment thresholds for fusion detection, but was impractical for screening a large cohort of samples or BAM files from suboptimal aligners in this context.

We therefore developed a focused automated algorithmic approach to evaluate NGS data specifically for integration events of TTMV within the *RARA* breakpoint cluster region around intron 2.^19,20^ The approach employs standard fusion detection principles based on chimeric split-reads and discordant reads, but also integrates alignments from different aligners to both the human genome and TTMV genomes. The code is available at github.com/ht50/ttmv_rara_SR (last accessed 2/21/2024) and addresses two main objectives:

1. The first objective was to efficiently screen for potential mutant TTMV::*RARA* reads, by using samtools (version 1.17) to extract a select subset of reads from an input NGS bam file and then applying blastn (BLAST+ version 2.13.0) to search for alignments to TTMV (NCBI taxonomy id 93678 in the nt_viruses database). If the input BAM file was generated by a modern local aligner and included supplementary and unmapped alignments to the human genome, then it was sufficient to extract the subset of paired-end reads harboring an alignment of either read of the pair to an approximate *RARA* breakpoint cluster region (chr17:38486109-38505716 in hg19 or chr17:40329857-40349464 in hg38, spanning *RARA* exon 2, intron 2, and exon 3 buffered by an additional 1 kb on each side). Otherwise, unmapped reads and their paired reads were also extracted from either the BAM file or the original FASTQ files. In pseudocode:

- Input: [bam] file including unmapped reads

- samtools view [bam] [region] | cut –f 1 | sort -u > [screen_reads]
- samtools view –f 4 [bam] | cut –f 1 | sort -u >> [screen_reads] (if needed)
- sort -u [screen_reads] -o [screen_reads] (if needed)
- samtools view –b –N [sceen_reads] [bam] > [screen_bam]
- samtools fasta [screen_bam] > [screen_fa]
- blastn –query [screen_fa] –db [nt_viruses] –taxids 93678 –outfmt 6 –out [ttmv.out]
- if [! -s ttmv_out]; then rm [screen_files]
2. The second objective was to evaluate reads found to have blastn alignments to TTMV, and to establish strong evidence of TTMV::*RARA* through identifying mutant junctional reads with chimeric split-read alignments including both (i) a blastn alignment of one part of the read to a TTMV accession number and (ii) a local alignment of another part of the read to *RARA*. The *RARA* local alignments were obtained from the intermediate BAM ([screen_bam]) when a modern local aligner was used to generate the input BAM, otherwise a realignment step was performed with bwa mem (version 0.7.17). For each chimeric alignment, mutant breakpoints were calculated and aggregated into counts according to TTMV accession number and by breakpoints. Non-chimeric discordant alignments, which were characterized by paired-end reads with only blastn alignments to TTMV for one read of the pair and only local alignments to *RARA* for the other read of the pair, were also counted but were not associated with mutant breakpoints. The TTMV accession number with the highest overall count from both chimeric and non-chimeric alignments was likely to be the best TTMV match.

A final objective was to assemble and characterize a TTMV::*RARA* contig or contigs. To this end, various assemblers including TRINITY^21^ (version 2.5.1) for RNA data and VELVET^22^ (version 1.2.10 using k-mer lengths of 15-29) for DNA data were applied to paired-end reads with a blastn alignment of either read of the pair to TTMV, supplemented by paired-end reads where at least one read of the pair had a partial local alignment to the approximate *RARA* breakpoint cluster region but without a supplementary, secondary, or primary alignment resolving the 5’ or 3’ part of the read. The resulting contigs were then aligned to TTMV by blastn and locally aligned to the human genome by bwa mem or blat in order to maximally characterize the fusion event. The contig with the highest blastn alignment score was also evaluated further by blastx to TTMV.

The algorithm was used to screen the BCH, MGH, and FMI cohorts. As an additional proof of concept, public RNA-sequencing data from the initial publication describing APL with *TTMV*:RARA was also re-analyzed by downloading FASTQ files from the NCBI sequence read archive (SRR14060870, SRR14060871), aligning to hg19 using bwa mem, and applying the algorithm.^10^ Software used in this project included applications installed and configured by BioGrids.^23^

### Anellovirus::*RARA* screening

To screen more widely across the anelloviridae family of viruses, the TTMV::*RARA* script included an option to replace the taxonomy id 93678 for TTMV with a fixed list of taxonomy ids from anelloviruses (github.com/ht50/ttmv_rara_SR/blob/main/anellovirus.txids). The list consisted of 645 taxonomy ids obtained by executing the blast script “get_species_taxids.sh –t 687329”, where 687329 is the taxonomy id for the family anelloviridae.

### Expressed TTMV::*RARA* quantification

In RNA-based fusion panel cases, analysis of *RARA* isoform junctions was performed as previously described to enable RNA quantification of TTMV::*RARA* in terms of an expressed variant allele frequency (VAF).^24^ In cases where TTMV was directly spliced to *RARA* exon 3, an expressed VAF of the fusion was calculated by counting TTMV::*RARA* split-reads spliced to exon 3 and dividing by split-reads of all junctions spliced to exon 3 derived from gene-specific primers targeting exon 3 or downstream exons. In cases where TTMV::*RARA* fusion transcripts connected TTMV to *RARA* intron 2 and also harbored aberrant cryptic *RARA* splicing connecting a segment of intron 2 to exon 3, empirical analysis was performed to confirm that the splice event did not occur in a historical cohort, and the expressed VAF of the cryptic isoform junction then served as a proxy quantification for the fusion. For serial samples that were subsequently negative for TTMV::*RARA* (based on the annellovirus::*RARA* algorithm, isoform split-read analysis, and manual review), an additional step of grep analysis of (i) the 30 bp junctional sequence spanning the diagnostic TTMV::*RARA* mutant junction and (ii) its reverse complement was also applied to the raw FASTQ files prior to UMI reduction, to further support an absence of the fusion.

## Results

### Case identification

We searched several clinical cohorts for APL with TTMV::*RARA*, resulting in detection of four cases. Both methods of custom analysis (manual review and automated algorithm) were capable of identifying TTMV::*RARA* fusions in all four cases, whereas the clinically validated pipelines of the RNA and DNA-based panels were consistently negative since these pipelines were not intended to detect fusions from viral genomes. The cases were discovered as follows. First, retrospective analysis of BCH pathology records identified five candidate acute leukemias from 2012-2022 with clinicopathologic features of APL but without *PML::RARA* or other driver fusions by conventional testing (karyotype, FISH, and/or RT-PCR). These five candidates were then tested by the fusion panel, and one of the five was confirmed to harbor TTMV::*RARA* fusion (Case 1) upon custom analysis. Using this experience, Case 2 was identified at BCH at the time of initial leukemia diagnosis, where clinical ordering of the fusion panel and custom analysis for TTMV::*RARA* were prompted by pathologist recognition of pathologic features suspicious for APL in the absence of detectable *RARA* fusion by conventional testing. Thus, identification of Case 1 and Case 2 required both (i) the decision to submit for fusion panel testing and (ii) custom analysis of the data. Manual review was the initial method of detection in Cases 1-2 (**Figure 1A-B**), followed by algorithmic analysis for enhanced characterization. The remainder of the BCH cohort and the entire MGH cohort was negative for TTMV::*RARA* by the algorithmic approach. Case 2 also underwent DNA panel sequencing within the FMI cohort, where the TTMV::*RARA* fusion on the DNA level was identified retrospectively by custom analysis. Cases 3 and 4 were then identified by application of the algorithmic approach to the remainder of the FMI cohort.

**Figure 1:**
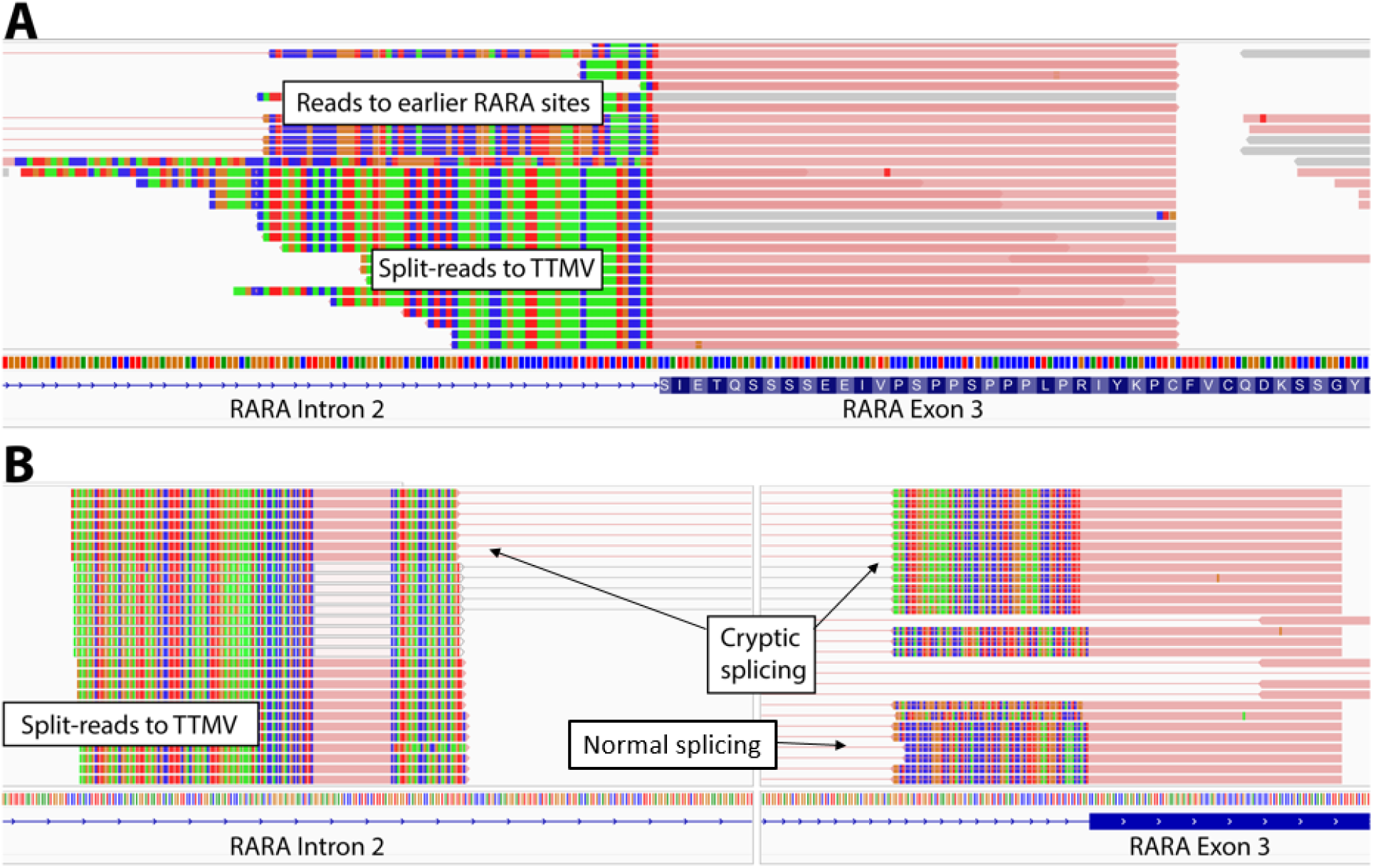
Manual review of Cases 1 and 2 in IGV. A) Case 1: Manual review in IGV of alignments over *RARA* exon 3 revealed split-reads with 3’ alignments to exon 3 and a novel 5’ soft clip pattern where the soft clips did not align to the human genome but were found to align by blastn to a TTMV isolate with high sequence identity. The primary split-read alignments in gray had unmapped paired reads that consisted predominantly or entirely of viral sequence. B) Case 2: Manual review in IGV of alignments within *RARA* intron 2 (left panel) and over exon 3 (right panel) revealed paired-end reads with primary and supplementary chimeric alignments consistent with aberrant cryptic splicing connecting a 26 bp segment within intron 2 (chr17: 38494366-38494391) to exon 3. Empirical analysis of historical fusion panel data showed this cryptic splice junction to be unique to this patient. The split-read alignments over the intron 2 segment moreover had 5’ soft clips (left panel) that did not align to the human genome but were found to align by blastn to several TTMV isolates with high sequence identity.

As an additional proof of principle, the algorithmic approach was applied to whole transcriptome RNA sequencing data from the inaugural TTMV::*RARA* publication, yielding *RARA*::TTMV::*RARA* contigs that recapitulated and refined previously described findings (**Supplementary Figures 1A-B**).^10^ Namely, the contigs demonstrated out-of-frame spliced insertions between *RARA* exons 2 and 3 of chimeric material (274 bp and 383 bp in length) composed of 5’ ends with high sequence identity to TTMV isolates and 3’ ends derived from short segments of *RARA* intron 2, thereby disrupting the standard frame of *RARA* but predicted to instead utilize a TTMV open reading frame (ORF) containing a putative conserved domain from the TT viral ORF2 superfamily and sharing the frame of *RARA* exon 3 (**Supplementary Figure 2A-B**). The fusion transcripts were thus inferred to contain *RARA* exons 1-2 but the encoded proteins were predicted to lack the portion of the N-terminal activation function domain corresponding to *RARA* exon 2. Of note, SRR14060871 also indicated cryptic splicing out of ∼1.7 kb within the TTMV component of its *RARA*::TTMV::*RARA* contig, thereby joining its ORF2 segment to a short, out-of-frame segment within ORF1 (**Supplementary Figure 2B**). Although TTMV::*RARA* breakpoints have usually been characterized within TTMV ORF2, this finding suggests that they may occur more widely throughout the TTMV genome, where splicing within TTMV and/or *RARA* consistently leads to joining of TTMV ORF2 to *RARA* exon 3 in the same frame.

### TTMV characteristics

For three of the four cases (Cases 2-4), the blastn alignments were predominantly to a collection of six highly homologous TTMV isolates in the viral database (**Table 1**), featuring pairwise identities of 95.50-99.82% relative to one another (**Supplementary Table 1**). By contrast, Case 1 had blastn alignments to only a single TTMV isolate that was considerably different, with pairwise identities of 74.35-74.65% relative to the six isolates from Cases 2-4. Similarly for the two cases from the original publication, the blastn alignments were predominantly to a collection of five highly homologous TTMV isolates, with pairwise identities of 96-100% relative to each other, 78.44-84.15% relative to the six isolates of Cases 2-4, and 77.32-78.12% relative to the isolate from Case 1.

**Table 1:** Number of TTMV::RARA split-reads (and discordant reads)

| GenBank_id | Case1_RNA | Case2_RNA | Case2_DNA | Case3_DNA | Case4_DNA | SRR14060870 | SRR14060871 |
| --- | --- | --- | --- | --- | --- | --- | --- |
| MN768572.1 | **67 (14)** |  |  |  |  |  |  |
| MN771921.1 |  | 4472 (1911) | 15 (590) | 8 (96) | **9 (209)** |  |  |
| MN774502.1 |  | 4472 (1911) | 15 (632) | 8 (99) | 9 (204) |  |  |
| MN771618.1 |  | 4472 (1911) | 15 (632) | **8 (99)** | 9 (204) |  |  |
| MN772671.1 |  | 4472 (1911) | 15 (632) | 8 (97) | 9 (204) |  |  |
| MN770942.1 |  | **4475 (2272)** | **15 (646)** | 4 (70) | 9 (204) |  |  |
| MN771042.1 |  | 4472 (1911) | 15 (632) | 4 (77) | 9 (204) |  |  |
| MN771954.1 |  | 895 (1163) |  |  |  |  |  |
| MN770247.1 |  | 2 (0) | 9 (101) |  |  |  |  |
| MN770240.1 |  | 2 (0) | 9 (101) |  |  |  |  |
| MN773107.1 |  |  | 9 (88) |  |  |  |  |
| MN773565.1 |  |  | 9 (88) |  |  |  |  |
| BK032512.1 |  |  | 9 (79) |  |  |  |  |
| MN772994.1 |  |  | 9 (88) |  |  |  |  |
| MN771972.1 |  |  | 9 (101) |  |  |  |  |
| MN770927.1 |  |  | 9 (101) |  |  |  |  |
| MN770954.1 |  |  | 9 (101) |  |  |  |  |
| MN770239.1 |  |  | 9 (101) |  |  |  |  |
| MN770241.1 |  |  | 9 (101) |  |  |  |  |
| MN770932.1 |  |  | 9 (101) |  |  |  |  |
| MN771787.1 |  |  |  |  |  | 355 (1269) | 110 (277) |
| MN774000.1 |  |  |  |  |  | 354 (1269) | 110 (277) |
| MN769771.1 |  |  |  |  |  | 354 (1269) | **110 (277)** |
| MN771920.1 |  |  |  |  |  | **354 (1273)** | 110 (277) |
| MN768731.1 |  |  |  |  |  | 351 (1266) | 110 (277) |
| MN772835.1 |  |  |  |  |  | 313 (1071) | 110 (276) |
| MN771783.1 |  |  |  |  |  | 71 (1260) | 97 (276) |

Shaded cells are the isolates for which the assembled contig had the highest blastn score. Of note, it generally was also associated with the highest number of blastn alignments. The split-reads and discordant reads are from alignments to TTMV isolates by blastn and to the human genome by bwa-mem.

Split-read analysis of RNA fusion panel data demonstrated TTMV spliced directly to *RARA* exon 3 (Case 1) or fused to a short segment (26 bp) within *RARA* intron 2 (chr17:38494366-38494391) followed by cryptic splicing to *RARA* exon 3 (Case 2) (**Supplementary Table 2**). The TTMV::*RARA* fusion junction of Case 2 (TTMV to chr17:38494366) represented expression of the underlying DNA breakpoint as confirmed later by the DNA panel, whereas in Case 1, the DNA breakpoint was spliced out on the RNA level but was predicted to occur similarly within *RARA* intron 2. RNA-based assembly generated chimeric contigs of lengths 413 bp (Case 1) and 574 bp (Case 2) whose first 204 bp and 337 bp aligned respectively to TTMV isolates MN768572.1 (98% identity) and MN770942.1 (96% identity) by blastn, and whose remaining 3’ ends aligned to *RARA* (100% identity) (**Figure 2A-B**). The blastn alignments indicated splicing out of short segments (87-90 bp) from TTMV in both Cases, followed immediately by potential start codons (ATG) whose open reading frames (ORF) were in the frame of *RARA* exon 3 and predicted by blastx to encode putative conserved domains from the TT viral ORF2 superfamily (pfam02957 amino acids 3-47; E-value < 1e-5) (**Figure 2C-D**). This splicing pattern was also observed within a predicted minor fusion isoform of SRR14060871 (**Supplementary Figure 2C**). The blastn alignments of Case 1 ended immediately before a GT dinucleotide sequence from TTMV (MN768572.1), consistent with a splice event to *RARA* exon 3.

**Figure 2:**
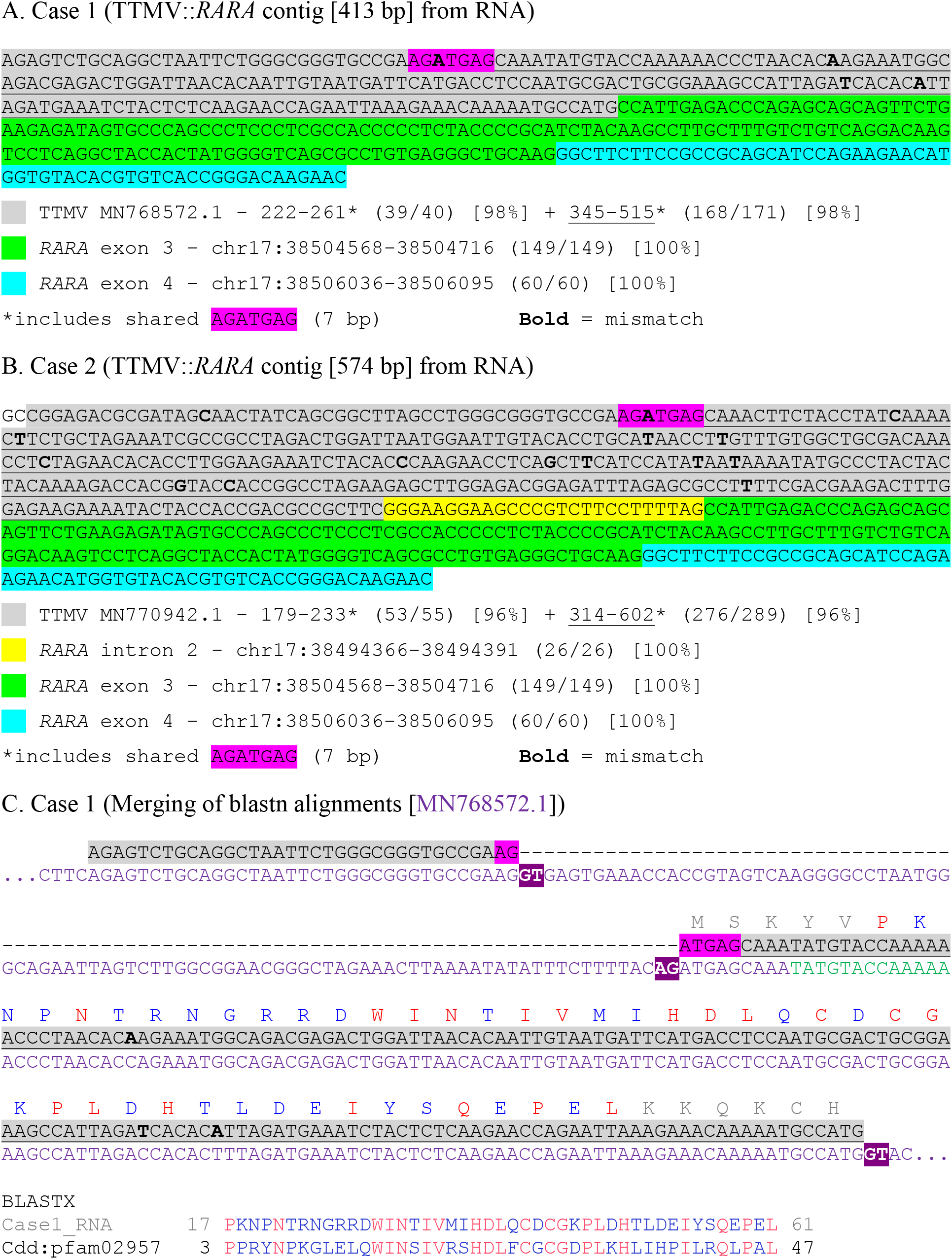
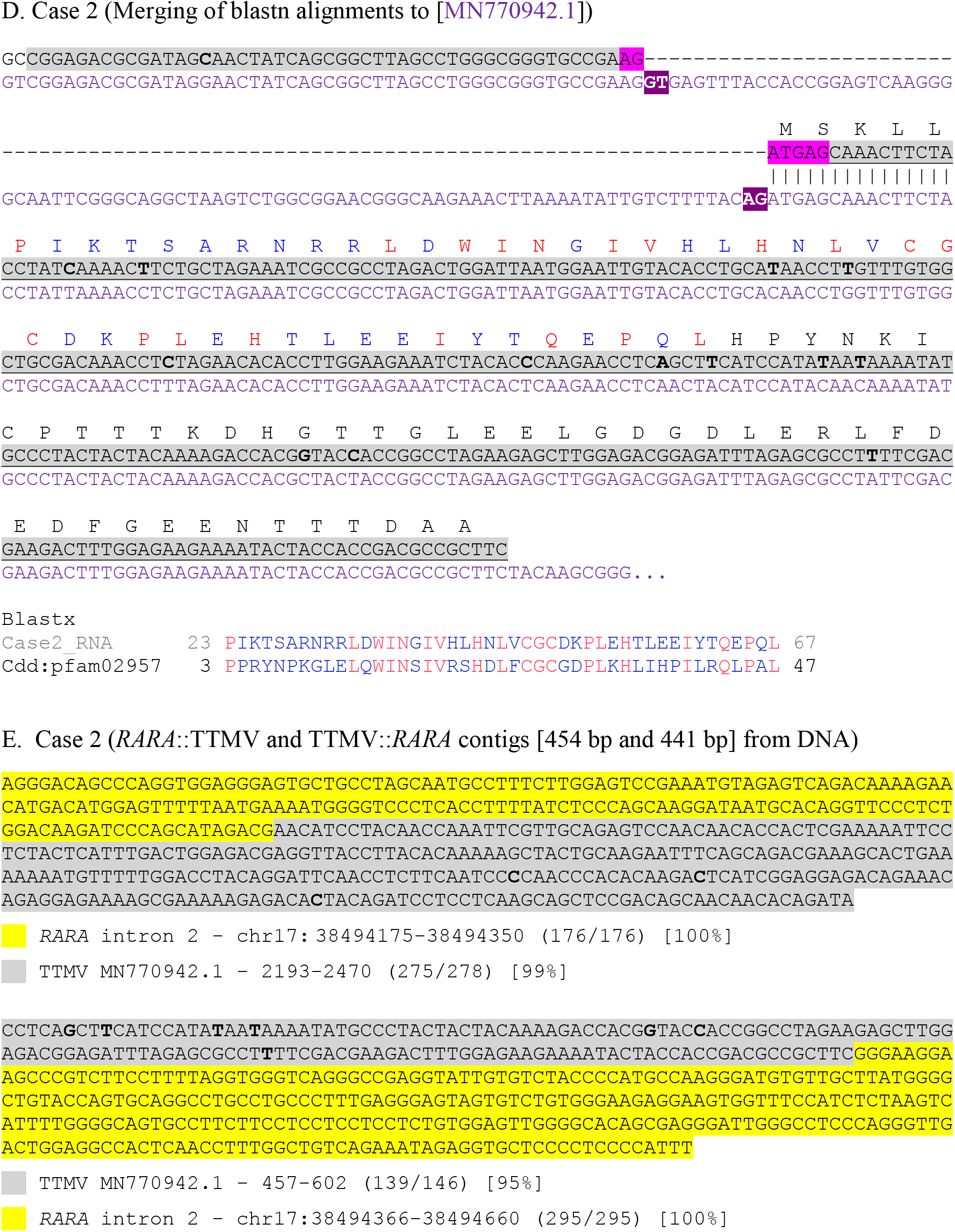
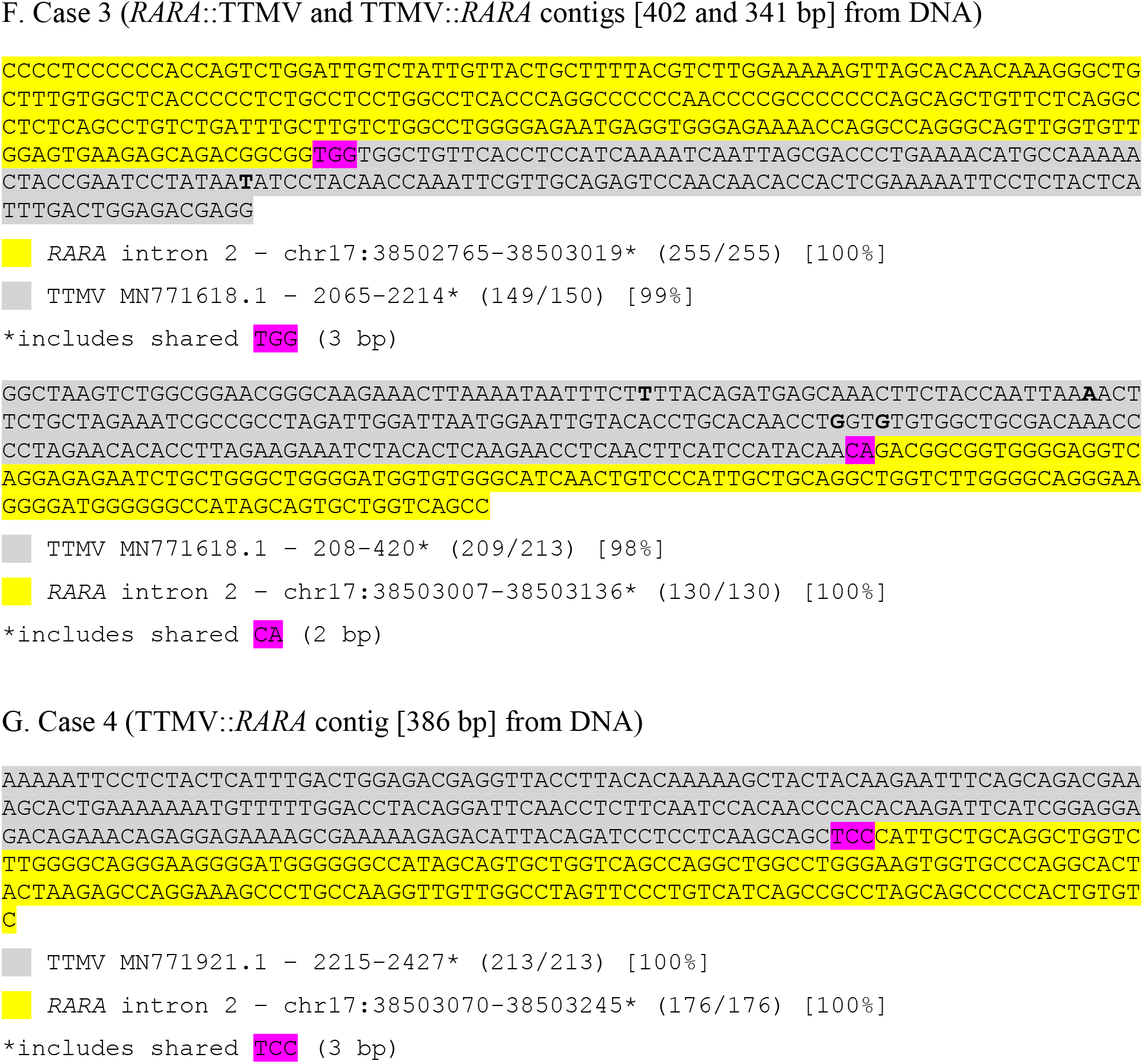
Fusion contigs of Cases 1-4. A-B) RNA-based contigs for Case 1 (A) and Case 2 (B) were generated from BCH fusion panel data using Trinity, yielding lengths of 413 bp and 574 bp. The 5’ ends of the contigs aligned best to the TTMV isolates MN768572.1 (Case 1; 96% identity) and MN770942.1 (Case 2; 98% identity), where both cases were characterized by 2 separate blastn alignments that overlapped in the same 7 bp sequence (AGATGAG). The remaining 3’ ends of the contigs aligned with 100% identity to regions of *RARA*: a short segment within intron 2 (Case 2 only), the entire exon 3 (Cases 1-2), and the initial 60 bp of exon 4 terminating at the end of an assay primer (Cases 1-2). C-D) Merging of the blastn alignments in both Case 1 (C) and Case 2 (D) was most consistent with splicing out of 87-90 bp segments from the respective TTMV genomes, where the inferred splice acceptor sites directly preceded potential ATG start codons of open reading frames that were predicted by blastx to encode putative conserved domains from the TT viral ORF2 superfamily pfam02957. E-G) DNA-based contigs for Case 2 (E), Case 3 (F), and Case 4 (G) were generated from FMI fusion panel data using Velvet, yielding *RARA*::TTMV and TTMV::*RARA* contigs for Case 2 (lengths 454 bp and 441 bp) and Case 3 (402 bp and 341 bp) but only a TTMV::*RARA* contig for Case 4 (386 bp). All *RARA* portions of the contigs aligned with 100% identity to regions within *RARA* intron 2, consistent with viral integration of TTMV within *RARA* intron 2. The TTMV::*RARA* breakpoints on the TTMV side were within ORF2 for Cases 2-3, similar to most previously characterized cases, and within ORF1 for Case 4, similar to findings from our re-analysis of SRR14060871 (see Supplementary Figures 1B and 2B).

Split-read analysis of DNA data demonstrated both *RARA*::TTMV and TTMV::*RARA* junctions in Case 2 and Case 3, where companion breakpoints were separated by only 16 bp (Case 2) and 12 bp (Case 3) within *RARA* intron 2 but were much further apart (estimated over 1 kb in both Cases) within TTMV, overall consistent with viral integration of linearized segments of TTMV within *RARA* intron 2 (**Supplementary Figure 3; Supplementary Table 2**). DNA-based assembly generated contigs of 441-454 bp (Case 2) and 341-402 bp (Case 3) surrounding the integration sites, however the DNA panel did not sequence across the complete viral insertion (**Figure 2E-F**). The predicted sizes of the viral insertions based on TTMV breakpoints were approximately 1257 bp (Case 2) and 1108 bp (Case 3). In Case 4, only a TTMV::*RARA* breakpoint (at chr17:38503070 within *RARA* intron 2) was detected and generated a 386 bp contig by assembly (**Figure 2G**), where the short read length size (49 bp) of the DNA panel may have reduced the sensitivity for detecting a companion *RARA*::TTMV breakpoint. Blastx of the assembled contig from Case 3 detected the same putative conserved domain from the TT viral ORF2 superfamily (pfam02957 amino acids 3-47; E-value < 1e-5) as Cases 1-2, whereas no conserved domain was detected in Case 4, however blastx yielded an alignment to TT viral ORF1 (pfam02956) of UGV36713.1 with 100% (68/68) identity on the protein level (**Supplementary Figure 4**). The DNA breakpoints connecting TTMV to *RARA* intron 2 thus occurred within ORF2 of TTMV in Cases 2-3, similar to most previously reported TTMV::*RARA* cases, and within ORF1 in Case 4, similar to our re-analysis of SRR14060871. RNA sequencing data was not available to characterize the transcription consequences on the RNA level for Cases 3-4, however the fusion transcripts were predicted to connect TTMV ORF2 in-frame to *RARA* exon 3 either directly or via cryptic splicing within TTMV and/or *RARA* intron 2, similar to prior cases characterized on the RNA level.

We also screened the BCH and MGH cohorts for anellovirus::*RARA* reads by running the *TTMV*::RARA script with the anellovirus option. No additional cases with split-reads or discordant reads between an anellovirus and *RARA* were found.

### Case descriptions (clinical course and pathologic features)

Clinical and pathologic features of the cases are summarized in **Tables 2 and 3**, respectively.

**Table 2:**
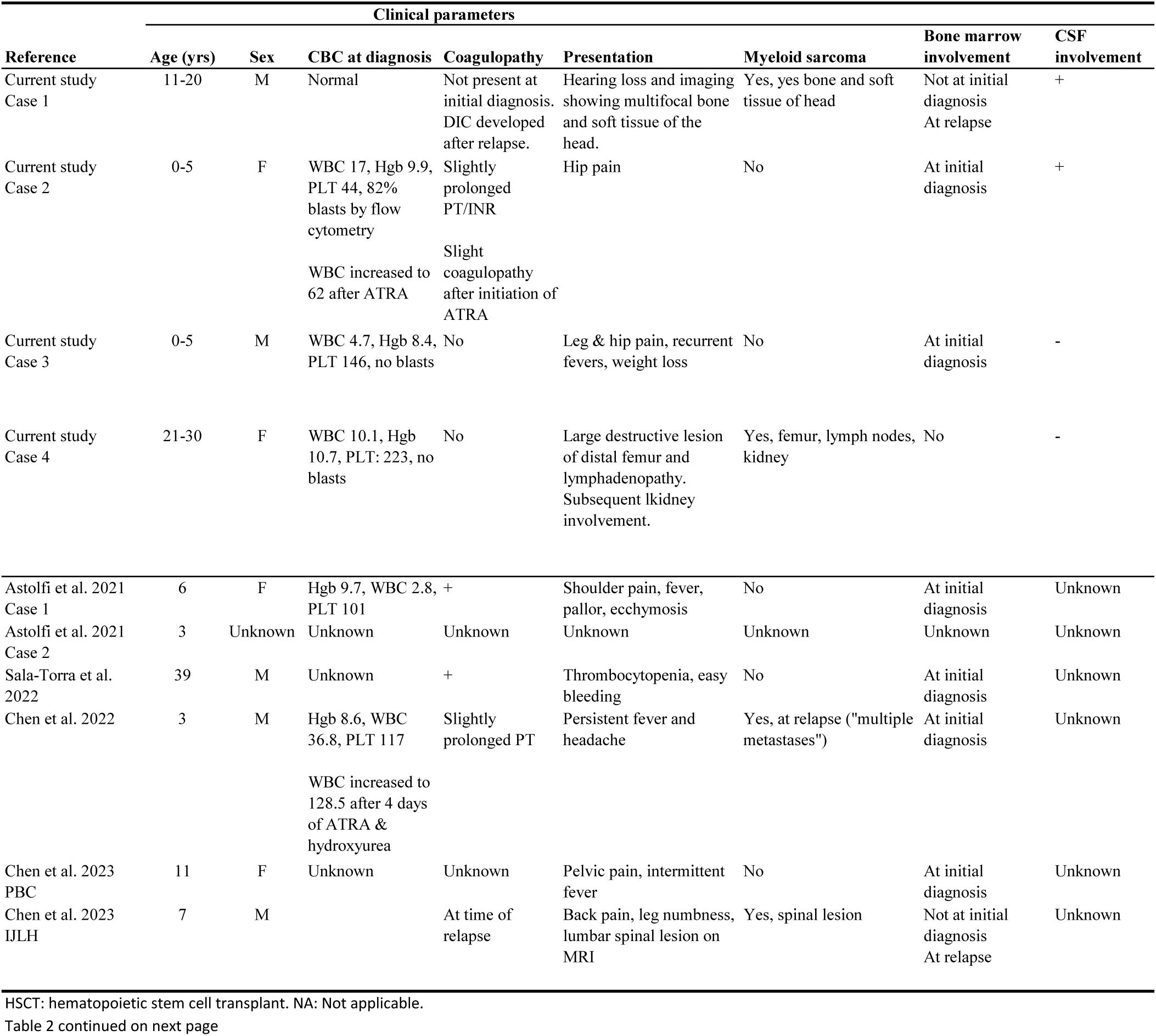
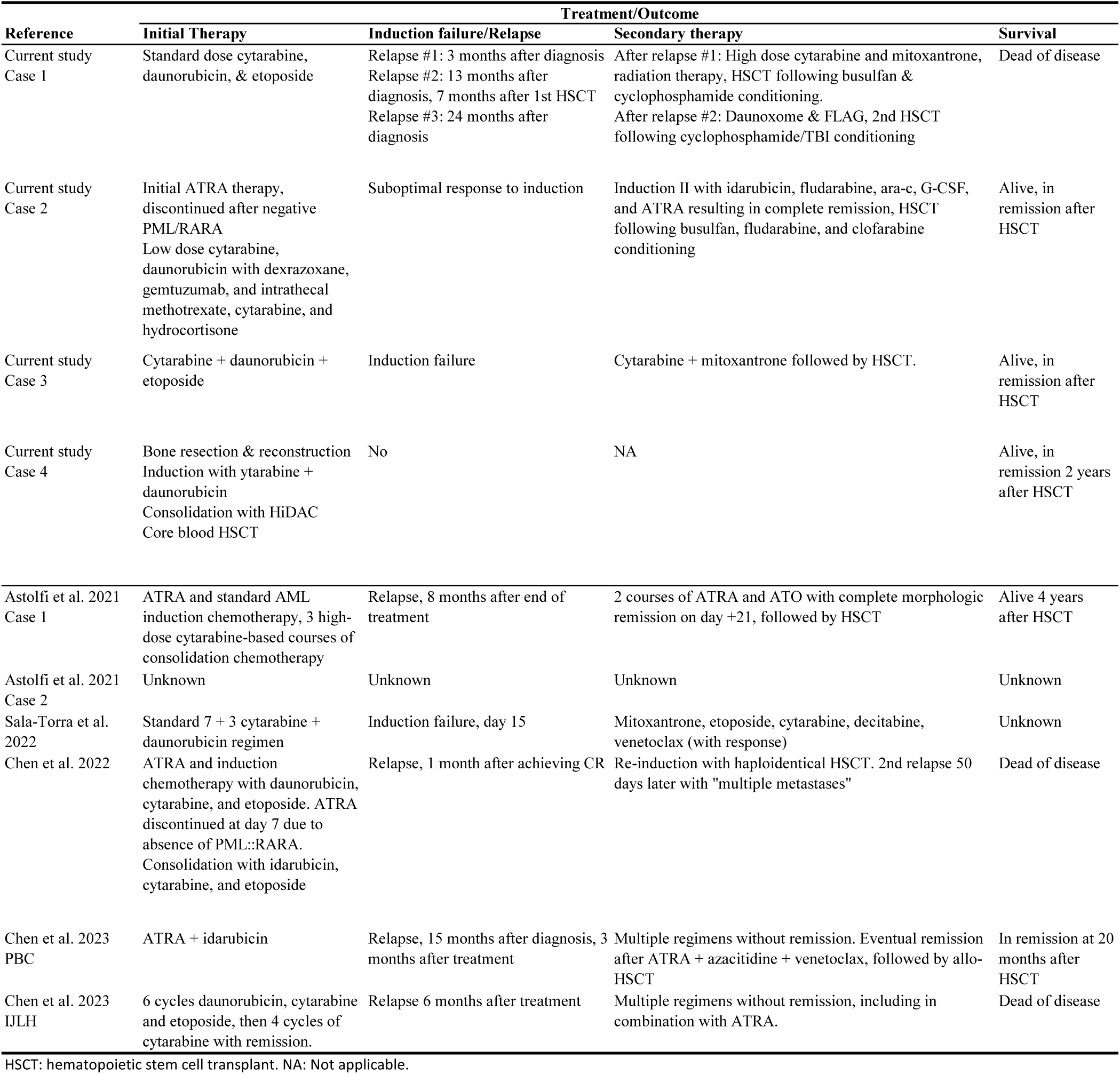
Clinical features and outcome of APL with TTMV::RARA fusion.

**Table 3:**
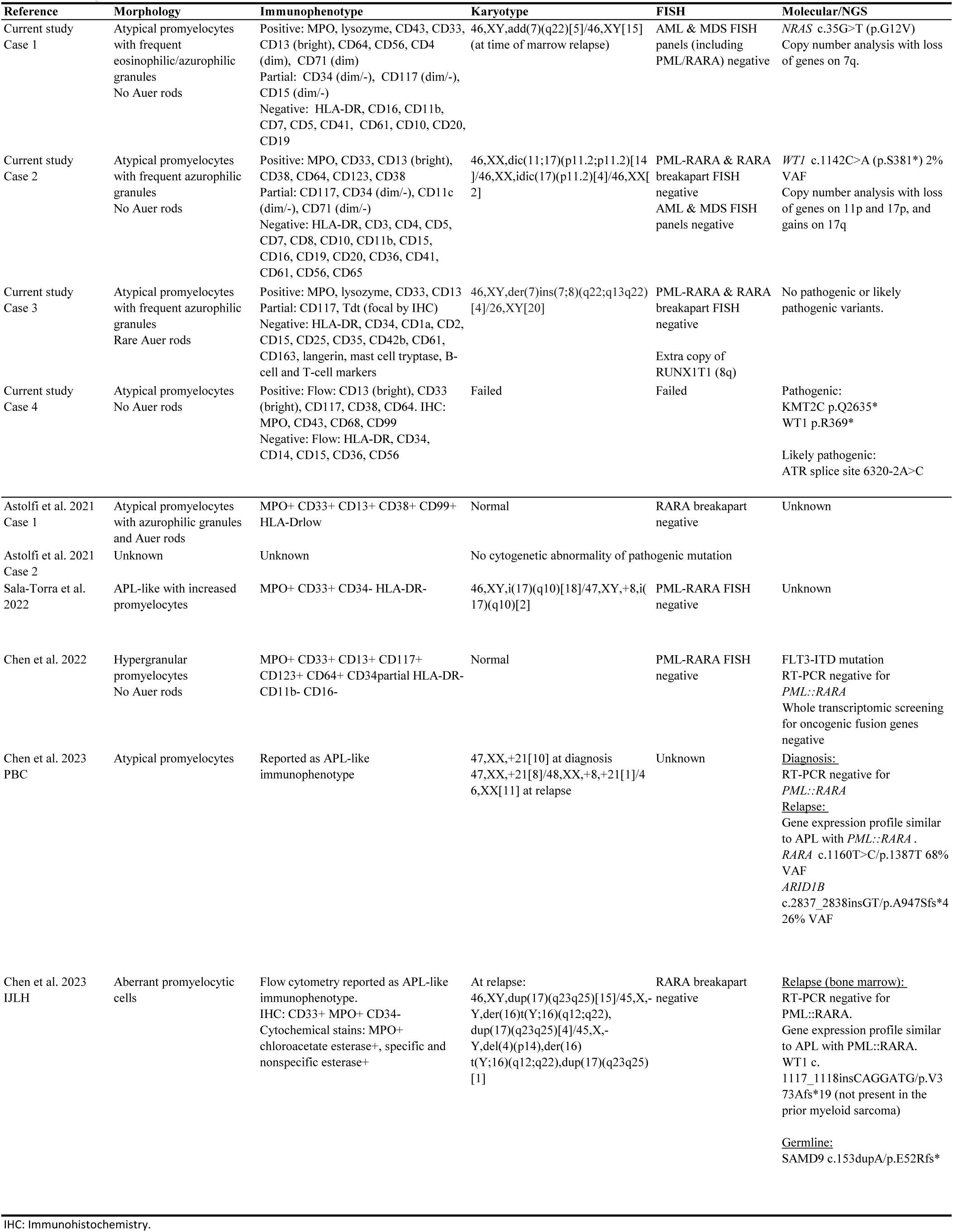
Pathologic features of leukemias with TTMV::*RARA*.

#### Case 1

An adolescent male (11-20 years old) presented with right-sided hearing loss and ear fullness. Imaging demonstrated destructive lesions involving the temporal bone, maxilla, and sphenoid wing. Complete blood count (CBC) and differential were normal. There was no evidence of coagulopathy. Specimens from surgical debridement demonstrated myeloid sarcoma (**Figure 3A**). The neoplastic cells had eosinophilic granular cytoplasm and ovoid to folded nuclei. Necrosis was present. By immunohistochemistry the neoplastic cells expressed CD43, MPO (**Figure 3B**), and lysozyme (subset), and were negative for CD34 (**Figure 3B inset**), CD117, CD56, and other markers. Dual-color/dual-fusion FISH was negative for *PML::RARA* rearrangement. Malignant cells were detected in the cerebrospinal fluid (CSF). The bone marrow was not involved.

**Figure 3:**
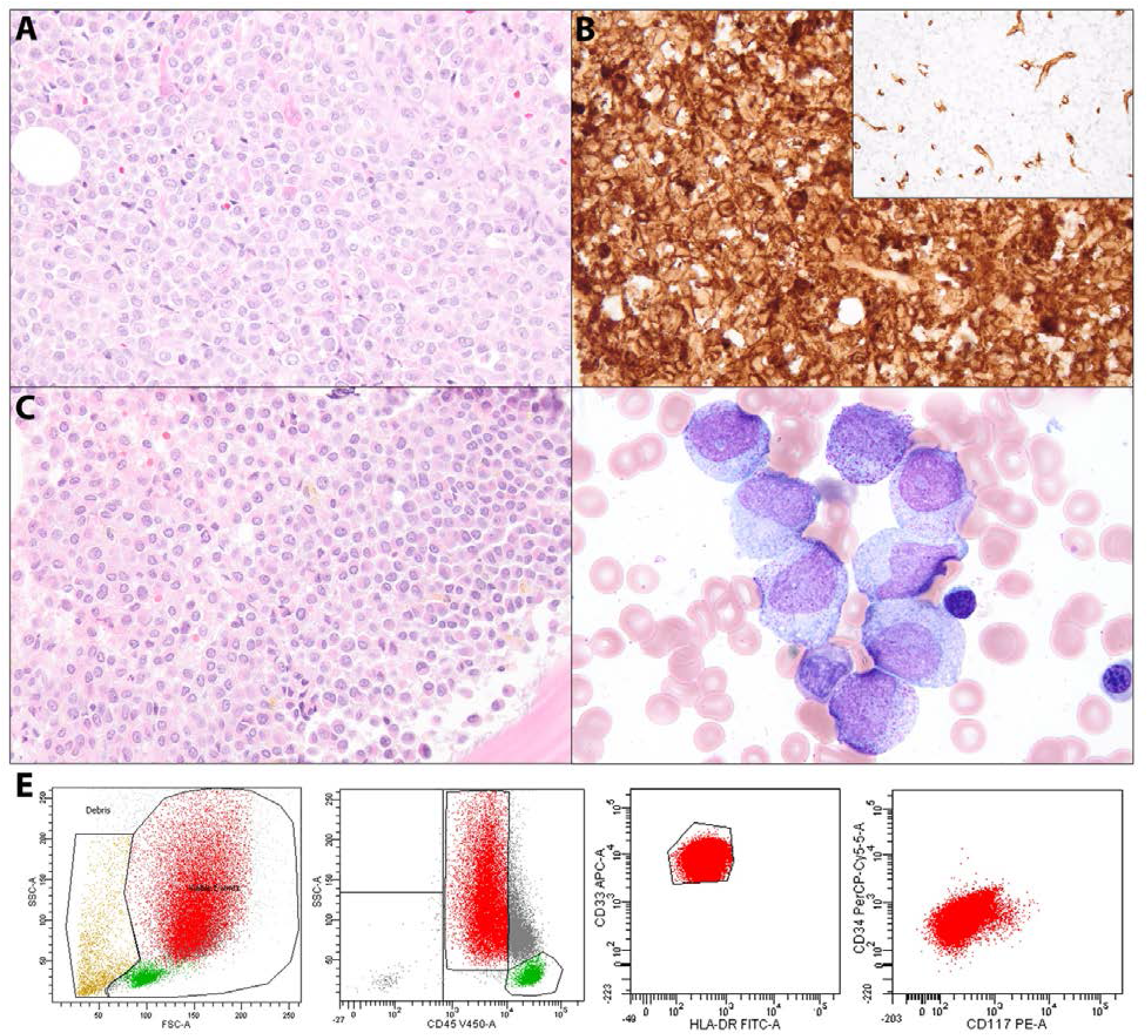
Pathologic features of Case 1. A-B) At initial diagnosis the debridement specimens demonstrated a sheet-like infiltrate of primitive cells with eosinophilic granular cytoplasm, ovoid to folded nuclei, dispersed chromatin, and variable small nucleoli, consistent with myeloid sarcoma (A). By immunohistochemistry the cells were positive for MPO (B) and negative for CD34 (B inset). C-E) Bone marrow involvement at the time of second relapse showed similar morphology with eosinophilic granular cytoplasm on the core biopsy (C). On the aspirate smear (D) the leukemic cells were large with ovoid to irregular nuclei, dispersed chromatin, variable pale nucleoli, and frequent azurophilic granules, including some heavily granulated forms. Flow cytometry (E) demonstrated that the leukemic cells had high forward and side scatter, dim CD45, and expressed CD33 with dim to negative HLA-DR, CD34, and CD117.

The patient underwent treatment with standard dose cytarabine, daunorubicin, and etoposide with weekly intrathecal therapy resulting in clearance of CSF leukemic cells. Three months after initial diagnosis, a paraspinal mass was incidentally noted on chest CT done for infection surveillance. PET/CT showed FDG-avid lesions involving the extra-pleural soft tissue, right lung base, and left iliac bone. Biopsy of the chest mass demonstrated relapsed myeloid sarcoma (relapse #1). Chromosome analysis and MDS/AML FISH panels were negative. The bone marrow was not involved.

The patient received high dose cytarabine and mitoxantrone and underwent radiation therapy to the right posterior mid-thorax, followed by conditioning with busulfan and cyclophosphamide and then HSCT. Seven months after HSCT the patient developed lymphadenopathy, cutaneous and subcutaneous soft tissue lesions involving the scalp. Biopsy of the scalp demonstrated relapsed myeloid sarcoma (relapse #2). Bone marrow biopsy demonstrated involvement by acute leukemia with promyelocytic features (**Figure 3C-D**). The leukemic cells were large with ovoid to indented nuclei, fine chromatin, and abundant cytoplasm with azurophilic granules. No definitive Auer rods were identified. Flow cytometry demonstrated leukemic cells with high SSC and expression of CD33, CD13 (bright), and MPO, with dim to negative CD34 and CD117, and negative for HLA-DR (**Figure 3E**). Chromosome analysis revealed an abnormal karyotype: 46,XY,add(7)(q22)[5]/46,XY[15]. An AML FISH panel including *PML::RARA* dual-color/dual-fusion FISH was negative. Given the absence of detectable *RARA* rearrangement or other driver, the leukemia was considered AML with maturation.

The patient underwent chemotherapy with liposomal daunorubicin and FLAG (fludarabine, cytarabine, and G-CSF) resulting in remission. He received a second HSCT following conditioning with cyclophosphamide/total body irradiation. Twenty-four months after initial diagnosis (7 months after 2^nd^ HSCT; 19 months after 1^st^ HSCT) the patient was found to have supraclavicular neck swelling. Biopsy demonstrated recurrent myeloid sarcoma (relapse #3) and the patient developed leukocytosis with peripheral leukemic cells and disseminated intravascular coagulation. He was transitioned to palliative measures and succumbed to progressive disease. Fusion panel testing was performed retrospectively on the myeloid sarcoma tissue and bone marrow specimens from the relapse #2 time point, with results described earlier.

#### Case 2

A previously healthy girl (0-5 years old) presented with hip pain and decreased activity. CBC demonstrated a WBC of 17 K cells/uL, anemia, and thrombocytopenia. PT and INR were slightly elevated. Leukemic cells with irregular nuclei, fine chromatin, prominent nuclei, and a moderate amount of cytoplasm containing azurophilic granules were present in the blood (**Figure 4A**) and bone marrow (**Figure 4B-D**). No Auer rods were identified. Flow cytometry demonstrated leukemic cells with increased SSC and expression of CD45(dim), MPO, CD13(bright), CD33, CD38, CD64, and CD117 (partial), with dim CD34 expression, and no expression of HLA-DR (**Table 3**; **Figure 4E**). A diagnosis of AML was rendered and suspicion for APL was raised based on morphology and immunophenotype. The CSF was involved. The patient received three doses of ATRA during which time she developed high fever, progressive leukocytosis and transient coagulopathy.

**Figure 4:**
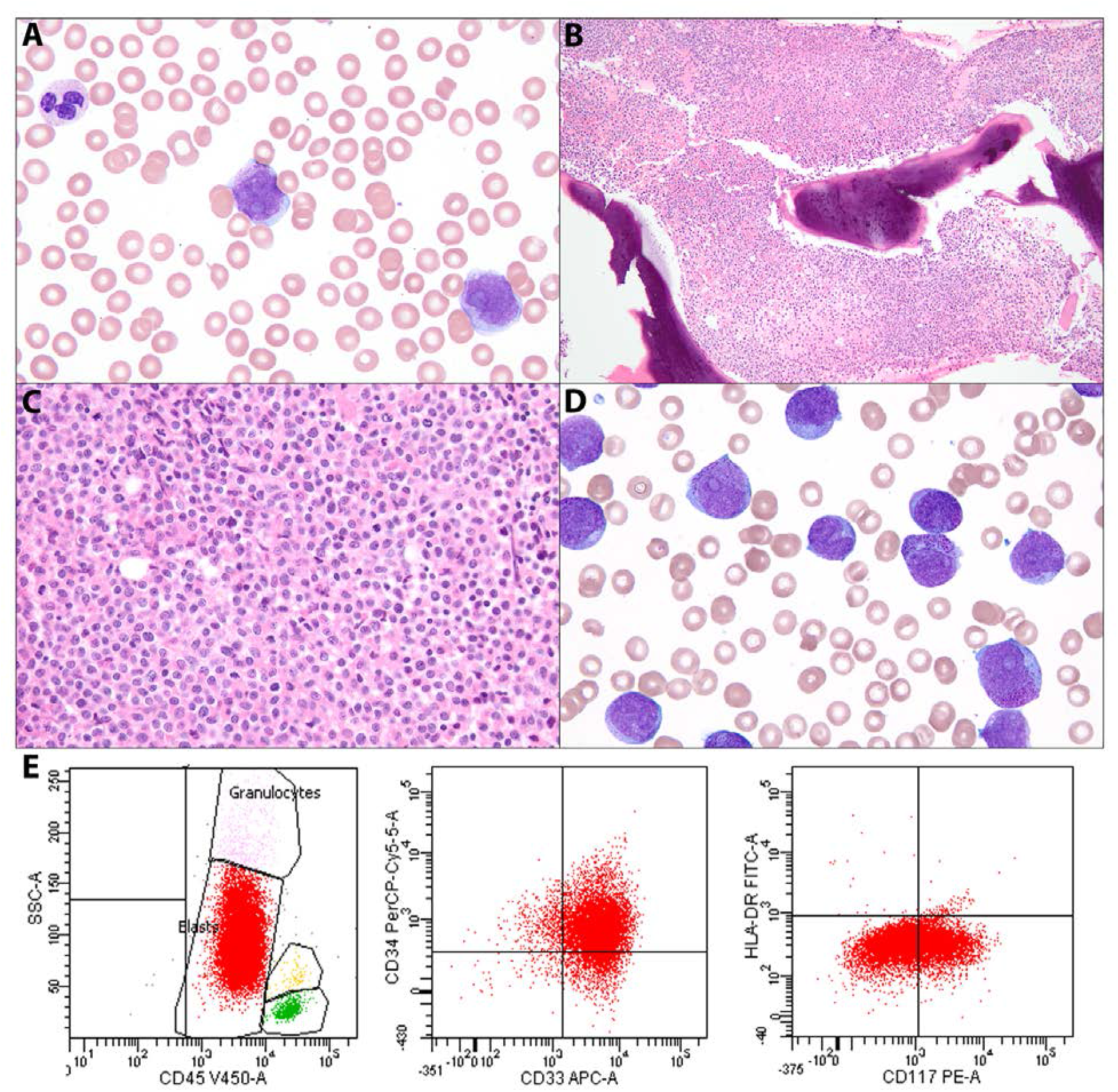
Pathologic features of Case 2. A) Peripheral blood smear demonstrated anemia, thrombocytopenia, and scattered large primitive myeloid cells with ovoid to irregular nuclei, dispersed chromatin, large nucleoli, and moderate cytoplasm with azurophilic granules. B-E) The bone marrow was involved by a sheet-like infiltrate of leukemic cells with eosinophilic granular cytoplasm and round to irregular nuclei on core biopsy (B-C). On aspirate smear (D) the leukemic cells were large with ovoid to folded nuclei, dispersed chromatin, variably prominent pale nucleoli, and frequent azurophilic granules, including some heavily granulated forms. Flow cytometry (E) demonstrated that the leukemic cells had increased side scatter and expressed CD45 (dim), CD34 (dim), CD33, CD117 (variable), with dim to negative HLA-DR.

*PML::RARA* dual-color/dual-fusion and *RARA* break-apart FISH analyses showed three non-rearranged copies of *RARA*. Chromosome analysis demonstrated an abnormal karyotype with two distinct clones, one with an unbalanced translocation between the short arms of chromosomes 11 and 17 resulting in deletion 11p and 17p, and the other with an isodicentric chromosome 17 resulting in trisomy 17q and monosomy 17p. Given the absence of detectable *RARA* translocation by cytogenetic studies, ATRA was discontinued and chemotherapy was initiated with standard dose cytarabine, daunorubicin with dexrazoxane, gemtuzumab ozogamicin, and intrathecal methotrexate, cytarabine, and hydrocortisone.

A routine DNA-based NGS assay (without *RARA*) demonstrated a pathogenic mutation in *WT1* c.1142C>A (p.S381*) at 2% variant allele frequency (VAF). Copy number analysis demonstrated one copy deletions of *WT1* on 11p and *PRPF8* and *TP53* on 17p, and partial gain of 17q, consistent with the karyotype.

In-house fusion panel testing demonstrated the cryptic TTMV::*RARA* fusion, as previously described. The leukemia was classified as APL with TTMV::*RARA*. Additional DNA-based NGS panel testing performed at FMI detected a pathogenic hotspot *TRRAP* p.S722F variant.

Bone marrow biopsy done after one course of induction chemotherapy showed residual acute leukemia at 3% by flow cytometry. Chromosome analysis showed a normal female karyotype. FISH demonstrated an extra non-rearranged copy of *RARA* in 2% of cells. Custom fusion analysis demonstrated persistent TTMV::*RARA* fusion at low level (see below). Given the suboptimal response, the patient proceeded to IDA-FLAG (idarubicin, fludarabine, cytarabine, and G-CSF) chemotherapy with the addition of ATRA on days 8-21 of the treatment cycle. Bone marrow biopsy done after the second treatment course demonstrated a flow cytometry negative remission. FISH and DNA-based NGS was negative for previously reported findings. Fusion panel testing showed a decreasing presence of the TTMV::*RARA* fusion to a very low level (see below). She then went on to receive high-dose cytarabine and etoposide plus ATRA for consolidation therapy. Again, she was flow, FISH and DNA NGS panel negative but continued to have the TTMV::*RARA* fusion identified at an extremely low level (see below).

The patient then received HSCT conditioned with busulfan, fludarabine, and clofarabine for disease consolidation. Bone marrow examinations done 35, 101, and 233 days later confirmed remission with no detectable TTMV::*RARA* fusion.

Fusion panel testing was used to track the TTMV::*RARA* fusion. At the time of initial diagnosis, the aberrant cryptic *RARA* splice junction (chr17:38,484,391 to chr17:38504568) connecting the 26 bp intron 2 segment to exon 3 within *TTMV::RARA* fusion transcripts had an expressed VAF of 80.5%. Subsequent fusion panel testing after treatment demonstrated expressed VAFs of 10.2% (after chemotherapy course one), 0.8% (after course two), 0.1% (after course three, pre-HSCT), and 0% (post-HSCT day +35, day +101, day +233).

#### Case 3

A previously healthy boy (0-5 years old) presented with recurrent fevers, weight loss, bilateral leg and hip pain, and recent viral infection. CBC demonstrated anemia and mild thrombocytopenia. LDH was elevated. Coagulopathy was present with elevated D-dimer and PT. MRI demonstrated marrow signal abnormality within the pelvis and proximal femur.

Bone marrow examination demonstrated a hypercellular myeloid-predominant marrow with 5% blasts and 19% promyelocytic cells along with maturing myeloid and erythroid elements. A differential diagnosis of early myeloid neoplasia versus regeneration after marrow insult was considered. Two weeks later thrombocytopenia worsened and 2% circulating blasts were reported. Repeat bone marrow examination demonstrated AML with features suggestive of APL including hypergranular promyelocytic cells with occasional Auer rods. The blasts expressed MPO, CD33, lysozyme, partial CD117, CD13(dim), and were negative for HLA-DR and CD34.

Cytogenetic analysis demonstrated an abnormal karyotype 46,XY,der(7)ins(7;8)(q22;q13q22) and FISH demonstrated an extra copy of *RUNX1T1* on 8q. NGS testing performed at FMI demonstrated no pathogenic or likely pathogenic variants. Fusion testing through the standard pipeline did not identify any fusions including *RARA*. However, retrospective custom analysis identified a TTMV::*RARA* fusion as described earlier.

The patient underwent AML induction chemotherapy with cytarabine, daunorubicin, and etoposide. Post-induction bone marrow showed persistent leukemia with 15% atypical promyelocytic cells. The patient underwent second induction with cytarabine and mitoxantrone and the subsequent bone marrow and ancillary studies confirmed remission. The patient underwent HSCT following conditioning with busulfan and cyclophosphamide, and all subsequent bone marrow examinations demonstrated continued remission. The patient is alive and disease free for >8 years. The CSF was negative for leukemic cells at all time points.

#### Case 4

A young adult woman (21-30 years old) presented with leg pain and imaging showed a large destructive lesion of the distal femur. CBC showed mild anemia only, with no circulating leukemic cells. There was no coagulopathy.

Several biopsies of the femur mass yielded only necrotic material. The patient underwent partial resection and reconstruction of the distal femur mass. The bone specimen and popliteal lymph node demonstrated myeloid sarcoma comprised of primitive large cells with open chromatin, indented nuclei, and eosinophilic granular cytoplasm, suggestive of promyelocytes or granular myeloblasts. Bone marrow iliac crest and CSF were negative for leukemia. Flow cytometry of the myeloid sarcoma demonstrated immature myeloid cells expressing CD13 (slightly bright), CD33 (bright), partial CD117, and CD38, but negative for CD34, HLA-DR, CD15, CD14, CD56, and CD10 (**Table 3**). By immunohistochemistry the lesional cells expressed CD45 (dim), MPO, CD43, and CD68.

FISH and cytogenetics were attempted but failed. NGS testing performed at FMI demonstrated pathogenic *KMT2C* p.Q2635* and *WT1* p.R369* variants, and a likely pathogenic *ATR* splice site variant c.6320-2A>C. Fusion testing through the standard pipeline did not identify any fusions including *RARA*. However, retrospective custom analysis identified a TTMV::*RARA* fusion, as described earlier.

PET/CT scan performed after 3 months demonstrated bilateral renal masses that were biopsied and confirmed to be myeloid sarcoma. The patient received AML induction chemotherapy with cytarabine and daunorubicin (7+ 3 regimen) with complete response, followed by consolidation with high dose intermittent cytarabine (HiDAC). The patient underwent HSCT and was known to be free of disease two years later.

## Discussion

We describe four cases of APL with TTMV::*RARA* fusion and provide methods to accurately identify and screen for the fusion, including clinicopathologic considerations and novel bioinformatics software that we have made publicly available. Like previously described cases, these share clinical, morphologic and immunophenotypic features of APL but without demonstrable *PML::RARA* rearrangement.^11^ This entity is relatively unknown among both clinicians and pathologists, but its recognition is important as it appears to be an aggressive disease and patients might benefit from incorporation of APL-directed treatment.^10^

The frequency of APL with TTMV::*RARA* is unknown. Given its only recent recognition and the few reports in the literature, this entity can be assumed to be rare, but is likely underrecognized.^10,12,13^ We retrospectively identified and tested five cases of AML with features of APL but without *PML::RARA,* of which one was found to harbor a TTMV*::RARA* fusion. Astolfi and colleagues retrospectively examined whole transcriptome sequencing data from 22 pediatric cytogenetically-normal AML cases and identified one with TTMV::*RARA* fusion.^10^ We screened fusion panel data from 50,257 additional cases of hematopoietic neoplasia including 782 pediatric and 4,027 adult AMLs and identified two cases. While selection bias cannot be excluded, these findings confirm that APL with TTMV::*RARA* is uncommon but appears to be enriched amongst leukemias with promyelocytic features, particularly within the pediatric and young adult age groups.

The cryptic nature of the TTMV::*RARA* fusion complicates diagnosis. Characteristics of TTMV insertions and TTMV::*RARA* fusions for reported cases are summarized in **Supplementary Table 3**. The inserted TTMV sequence size has been reported to range from an estimated 1045-2872 base pairs, which is below the level of resolution of standard cytogenetic techniques and thus undetectable by karyotype and FISH.^10,12,13^ Moreover, since the fusion involves a virus external to the human genome, molecular NGS fusion platforms may not detect TTMV::*RARA* fusions in their standard pipelines, which typically align sequencing data only to the human genome or occasionally to a limited set of viral genomes from established oncoviruses unlikely to include TTMV. Therefore, a high degree of suspicion based on clinicopathologic features is needed to prompt both testing that can robustly capture the TTMV::*RARA* fusion and custom analysis to recognize the fusion within the raw or intermediate data. Screening strategies include fusion/transcriptomic analysis of cases of AML with promyelocytic features or of any AML without a demonstrable molecular driver, followed by manual review of exon 3/intron 2 of *RARA,* or implementation of automated methods to identify such cases.

Knowledge of characteristic clinicopathologic features may prompt recognition of APL with TTMV::*RARA*. Eight of ten reported cases (including this study) have been children and adolescents, along with two adults (both under 40 years old).^10,12,13,25^ Coagulopathy was present in several cases, similar to APL with *PML::RARA*.^10,12,13^ CSF involvement, which is uncommon in APL with *PML::RARA*, was present in two of four cases presented here. After treatment with ATRA, two cases demonstrated a marked increase in WBC along with coagulopathy in one case.^13^ Myeloid sarcoma is frequent, occurring in four of the nine cases with clinical data, and was the initial presentation in three. In patients presenting with myeloid sarcoma, routine bone marrow examination was frequently negative for leukemia at the time of initial diagnosis but became positive after relapse.^13,15^ Characteristic leukemic cell morphology with frequent azurophilic granules and/or cells resembling promyelocytes were present in all described cases.^10,12–15,25^ Auer rods were present in only some cases.^13^ The leukemic immunophenotype was similar to APL with *PML::RARA* including absent or only partial/dim expression of HLA-DR and CD34 in all described cases, along with expression of MPO and other myeloid markers including CD33 and CD13.^10,12,13^ By flow cytometry, CD64 and bright CD13 expression were frequently present and may be characteristic.^13^

Isochromosome 17q and isodicentric 17p11 are uncommon cytogenetic abnormalities in AML.^23^ In adults, myeloid neoplasms with isolated iso(17q) typically lack *TP53* mutations and are characterized by mutations in *ASXL1*, *SRSF2*, and *SETBP1*, suggesting different pressures and mechanisms in formation of iso(17q).^26^ APL infrequently demonstrates iso(17q) or idic(17p11), where it usually co-occurs with t(15;17) resulting in an additional copy of *PML::RARA* but can also be an isolated finding associated with a cryptic *PML::RARA*.^27–29^ Interestingly, two of the described cases of APL with TTMV::*RARA* fusion demonstrate iso(17q) or idic(17p), suggesting that this may represent a characteristic chromosomal abnormality. Indeed, several cases of AML with promyelocytic features, iso(17q), and no detectable *PML::RARA* have been described, possibly representing APL with unidentified TTMV::*RARA* fusions.^30,31^

Three cases of APL with TTMV::*RARA* harbored *WT1* mutations, which is known to be enriched in APL with *PML::RARA* and variant fusion partners.^15,32,33^ *FLT3*-ITD and mutations in *NRAS*, *ARID1B,* and *KMT2C* have also been documented. *KMT2C* mutations appear to be relatively common in APL with variant fusions, whereas *FLT3*-ITDs and to a lesser extent *NRAS* mutations are known to be relatively frequent in APL with *PML::RARA*. Interestingly, one case reported by Chen et al. developed a mutation in the ligand-binding domain of *RARA* after therapy with ATRA suggesting leukemic clonal evolution to bypass ATRA efficacy.^14^ The same group reported another case in which germline frameshift *SAMD9* mutation was identified suggesting the possibility of germline predisposition.^15^ Gene expression profiling of several prior cases showed clustering of TTMV::*RARA* cases with the group of APL with *PML::RARA* and away from other AML.^14,15,25^

Based on the limited number of cases published to date, APL with TTMV::*RARA* appears to have an aggressive clinical course and poor response to standard AML chemotherapy. Of the nine cases with available clinical data, relapse occurred in five and induction failure or poor response to induction in three others. The appropriate treatment of APL with TTMV::*RARA* remains uncertain, but the presence of *RARA* translocation provides rationale for the use of APL-type therapy. Indeed, use of ATRA along with other agents resulted in complete remission in several cases including case 2 of this report.^10,1414^ ATRA has dual therapeutic effects in APL, it activates transcription of differentiation genes and leads to degradation of the PML-RARA oncoprotein.^34,35^ APL with certain *RARA* fusion partners are not sensitive to ATRA therapy.^32^ The emergence of synthetic retinoids with enhanced differentiation activity, like tamibarotene, has shown promise in rare fusions resistant to ATRA. ATO hypothetically has no therapeutic potential in APL with TTMV::*RARA* because its effect is mediated by its binding to PML of PML-RARA. This underscores the ongoing evolution of molecular testing on therapeutic strategies in APL.^36,37^

TTMV insertion characteristics are similar between all described cases. The TTMV DNA insertion site is within intron 2 and results in either direct splicing of TTMV ORF2 in-frame to *RARA* exon 3 (e.g. Case 1) or retention of a small segment of intron 2 separating the TTMV ORF2 sequence in-frame from exon 3 (e.g. Case 2). The breakpoints within TTMV usually occur within ORF2 but can also occur within ORF1 (Case 4), where reanalysis of public data (SRR14060870) showed how cryptic splicing within both TTMV and *RARA* connects ORF2 in-frame to *RARA* exon 3 with retention of small intervening segments of TTMV ORF1 and *RARA* intron 2. A previously reported case described an absence on the RNA level of 84-bp of viral sequence that was present in the inserted DNA.^12^ Similarly, our cases with RNA data showed blastn alignments with high sequence homology to TTMV isolates and predicted splicing out of homologous 87-90 bp segments of viral sequence.

The methods presented here for detecting TTMV::*RARA* in NGS data depended on the collection of TTMV isolates deposited to the NCBI viral database and the ability to generate blastn alignments that scored sufficiently high under the default settings. The blast database used at the time of this study included 6686 TTMV accessions, where the contigs from Cases 1-4 had up to 95.7-100% identities to 4 specific TTMV isolates respectively, of which 3 were highly homologous to one another. Similarly, contigs generated from public RNA sequencing data had 97.6% (203/208) and 100% (285/285) identity to the highly homologous TTMV isolates MN771920.1 and MN769771.1, while the other 2 cases described in the literature were reported to have 93-94% sequence identity to MN769771.1 once again,^12^ and to unspecified isolates.^13^ The recurrence of highly homologous TTMV isolates is notable but could have many different explanations, ranging from spatiotemporal patterns of TTMV infection to potential biological differences between isolates. Overall, the very high identities of all known TTMV::*RARA* cases to date is reassuring for effectively detecting TTMV::*RARA* by blast-based molecular methods. However, these methods theoretically could miss a TTMV::*RARA* fusion arising from a TTMV isolate that has not been submitted to, and is different from, the isolates in the viral database. Thus, the development of other algorithmic approaches may be worth further investment and might increase the yield of screening efforts. Such enhancements could include experimentation with blastn settings and exploration of blastx. Finally, an orthogonal method might be based on screening for alignments with softclips starting at a common genomic location within *RARA* intron 2 to exon 3 but without supplementary alignments resolving the softclip; however, this approach may be challenging to automate from the perspective of specificity.

Several methods have been used to identify TTMV::*RARA* fusions, including whole transcriptome or genome sequencing and long-range DNA sequencing with nanopore and CRISPR guides targeting *RARA*, typically combined with manual review of data. We demonstrate that the fusion is similarly detectable by manual analysis of fusion data from a clinical Archer-based RNA fusion assay, which is a commercially available platform used by many diagnostic molecular laboratories. Moreover, we develop an efficient automated method to eliminate the need for labor-intensive manual review and for more complete, routine evaluation of the data. We also show how the Archer assay enables effective monitoring of disease burden during treatment. The Archer assay should be capable of detecting any TTMV::*RARA* fusion with an RNA breakpoint that is not too distant from *RARA* exon 3, which has been the case for all previously described cases in the literature since the retained segments of intron 2 on the RNA level have always been relatively small. Of note, fusion panel assays without the capability to detect unknown *RARA* partners, without specialized informatics, or without appropriate manual analysis may not identify the fusion. The ability to detect the TTMV::*RARA* fusion relatively rapidly using the Archer-based clinical assay may be advantageous in comparison to less widely-available and more time-intensive testing modalities. Since the assay will also detect a wide range of fusions, isoforms, and expressed variants with diagnostic, prognostic, and therapeutic relevance, it may be useful and reasonable to order this test early in a clinical workup.

## Conclusion

We characterize four cases of APL with TTMV::*RARA* and summarize the clinicopathologic and molecular features of the cases described to date. We highlight characteristic features that should prompt clinician and pathologist recognition of this newly-described entity. We also provide a framework for how to identify and screen for the fusion by manual review of fusion panel data and automated analysis, and we demonstrate that the TTMV::*RARA* fusion may be otherwise cryptic on routinely-used diagnostic modalities including karyotype, *PML::RARA* fusion and *RARA* break-apart FISH. Determination of the true frequency of APL with TTMV::*RARA* will require larger studies, however our limited data suggests that it may be particularly rare in adults and enriched in pediatric leukemias with an APL phenotype. Future studies will be informative to characterize this entity in general and the biology of the TTMV insertions in particular, to identify more rapid means of fusion detection, and to investigate optimal therapeutic strategies.

## Data Availability

Code for the automated custom TTMV::RARA analysis described in this paper is available at: github.com/ht50/ttmv_rara_SR. For other data please contact the corresponding author.

## Contributing author contact information

HT

MS

MM

EW

AA

DRB

EMH

FW

LG

FL

VN

AP

AK

CB

BD

JP

MH

