## Supplementary Data for "Acute promyelocytic leukemia with torque teno mini virus (TTMV)::*RARA* fusion: an approach to screening and diagnosis"

**Supplementary Table 1: Pairwise BLASTN scores (and percent identities)**

| | MN771921.1 | MN774502.1 | MN771618.1 | MN772671.1 | MN770942.1 | MN771042.1 | MN768572.1 | MN771787.1 | MN774000.1 | MN769771.1 | MN771920.1 | MN768731.1 |
|---|---|---|---|---|---|---|---|---|---|---|---|---|
| MN771921.1 | 5193 (100%) | | | | | | | | | | | |
| MN774502.1 | 5012 (99.74%) | 5051 (100%) | | | | | | | | | | |
| MN771618.1 | 5055 (99.82%) | 5003 (99.78%) | 5083 (100%) | | | | | | | | | |
| MN772671.1 | 4891 (99.77%) | 4802 (99.66%) | 4859 (99.74%) | 4924 (100%) | | | | | | | | |
| MN770942.1 | 4532 (95.77%) | 4386 (95.61%) | 4418 (95.64%) | 4259 (95.50%) | 5258 (100%) | | | | | | | |
| MN771042.1 | 5153 (99.75%) | 5005 (99.71%) | 5048 (99.78%) | 4883 (99.74%) | 4635 (95.68%) | 5332 (100%) | | | | | | |
| MN768572.1 | 222 (74.48%) | 191 (74.35%) | 206 (74.44%) | 211 (74.55%) | 224 (74.65%) | 277 (74.48%) | 5230 (100%) | | | | | |
| MN771787.1 | 342 (81.78%) | 333 (84.15%) | 337 (82.90%) | 342 (83.16%) | 359 (78.44%) | 372 (82.22%) | 158 (77.32%) | 5402 (100%) | | | | |
| MN774000.1 | 350 (79.16%) | 340 (79.67%) | 344 (78.97%) | 350 (79.16%) | 370 (78.71%) | 372 (81.60%) | 148 (78.12%) | 5042 (96%) | 5241 (100%) | | | |
| MN769771.1 | 350 (79.16%) | 340 (79.67%) | 344 (78.97%) | 350 (79.16%) | 370 (78.71%) | 426 (81.60%) | 148 (78.12%) | 5230 (100%) | 5192 (100%) | 5494 (100%) | | |
| MN771920.1 | 355 (79.35%) | 346 (79.88%) | 350 (79.16%) | 355 (79.35%) | 375 (78.88%) | 377 (81.82%) | 148 (78.12%) | 5192 (100%) | 5158 (99.58%) | 5473 (99.62%) | 5520 (100%) | |
| MN768731.1 | 348 (82.71%) | 339 (83.75%) | 342 (82.46%) | 348 (82.71%) | 364 (78.53%) | 374 (78.74%) | 148 (78.12%) | 5042 (96%) | 5147 (99.40%) | 5177 (99.58%) | 5120 (99.33%) | 5243 (100%) |

**Supplementary Table 2: Split-read (SR) breakpoints**

| Sample | Best match | NGS | RARA::TTMV junction | #SR | TTMV::RARA junction | #SR |
|---|---|---|---|---|---|---|
| Case1 | MN768572.1 | RNA | | | ttmv:515_RARA:e3 | 67 |
| Case2 | MN770942.1 | RNA | | | ttmv:602_17:38494366 | 4366 |
| | | DNA | 17:38494350_ttmv:2193 | 9 | ttmv:602_17:38494366 | 6 |
| Case3 | MN771618.1 | DNA | 17:38503019_ttmv:2065_ov3 | 4 | ttmv:420_17:38503007_ov2 | 4 |
| Case4 | MN771921.1 | DNA | | | ttmv:2427_17:38503070_ov3 | 9 |
| SRR14060870 | MN771920.1 | RNA | RARA:e2_ttmv:492 | 289 | ttmv:699_chr17:38490465_ov-1 | 47 |
| | | | RARA:e2_ttmv:340 | 1 | | |
| | | | 17:38490448_ttmv:2629_ov4 | 10 | | |
| SRR14060871 | MN769771.1 | RNA | RARA:e2_ttmv:438 | 16 | ttmv:2502_17:38489987_ov2 | 88 |
| | | | 17:38490031_ttmv:2606_ov4 | 1 | ttmv:721_RARA:e3 | 4 |

ov2, ov3, ov4: ambiguous breakpoints due to overlapping alignments from microhomology of 2bp, 3bp, 4bp
ov-1: intervening unaligned nucleotide (1 bp)

**Supplementary Table 3: Characteristics of insertion and TTMV in leukemias with TTMV*::RARA* fusion**

| Reference | Method of TTMV::RARA detection | TTMV insertion site (hg19) and consequence in RNA transcripts | TTMV insertion/expressed sequence size (predicted) | Closest TTMV isolate | Percentage identity | Predicted protein size (from using TTMV ORF2) |
|---|---|---|---|---|---|---|
| Current study Case 1 | Manual review of Archer fusion analysis data followed later by | Unknown (predicted in intron 2) | Unknown, 169 bp from TTMV ORF expressed in fusion transcript | MN768572.1 MAG: Torque teno mini virus isolate SAfiA-656-10, complete ORF1 | 99% identity, unknown coverage | 459 amino acids |
| Current study Case 2 | Manual review of Archer fusion analysis data followed later by | Chr17:38494350; Chr17:38494366. Upstream of RARA exon 3 (in frame), linked by 26 nucleotides of intron 2 | Estimated 1257 bp insertion, 287-bp from TTMV ORF expressed in fusion transcript | MN770942.1 MAG: TTV-like mini virus isolate SAfiA-468-10, complete genome | 98% identity, 34% estimated coverage | 507 amino acids |
| Current study Case 3 | Automated algorithm analysis | Chr17:38503019; Chr17:38503007 with 2-3 bp of microhomology; unknown transcript | Estimated 1108 bp insertion | MN771618.1 MAG: TTV-like mini virus isolate SAfiA-235-53, partial genome | 99% identitiy, 33% estimated coverage | Unknown |
| Current study Case 4 | Automated algorithm analysis | Chr17:38503070 with 3bp of microhomology; unknown transcript | Unknown | MN771921.1 MAG: TTV-like mini virus isolate SAfiA-657-34, partial genome | 100% identitiy, unknown coverage | Unknown |
| Astolfi et al.[10] Case 1 | Whole transcriptomic sequencing | Chr17:38490448. Upstream of RARA exon 3 (in-frame), linked by 38 nucleotides of intron 2 | Estimated 1045 bp insertion, 209 bp expressed in fusion transcript | MN771920.1 MAG: TTV-like mini virus isolate SAfiA-652-8, partial genome | 98% identity, unknown coverage | 485 amino acids |
| Astolfi et al.[10] Case 2 | Retrospective in silico analysis in a whole transcriptomic sequencing database | Chr17:38490031. Upstream of RARA exon 3 (in frame), linked by 45 nucleotides of intron 2 | Estimated 2872 bp insertion, 328 bp expressed in fusion transcript | MN769771.1 MAG: TTV-like mini virus isolate SAfiA-858-85, complete genome | 100% identity, unknown coverage | 427 amino acids |
| Sala-Torra et al.[12] | Long-range sequencing with nanopore and CRISPR guides targeting RARA | Chr17:38494342. Upstream of RARA exon 3, linked by 14 nucleotides of intron 2 | 2450 bp insertion, up to 498 bp expressed in fusion transcript | MN769771.1 MAG: TTV-like mini virus isolate SAfiA-858-85, complete genome | 93% identity, 99% coverage | NA |
| Chen et al. 2022.[13] | Retrospective cluster analysis followed by manual investigation of RARA in whole transcriptomic sequencing data and whole genome sequencing data | Chr17:38488517. Upstream of RARA exon 3 (in-frame), within intron 2, resulting in direct splicing of TTMV to RARA exon 3 | 1163 bp insertion, 314 bp expressed in fusion transcript | NA | 93-94% identity, 83% coverage | 488 amino acids |
| Chen et al. 2023.[14] | Retrospective cluster analysis followed by manual investigation of RARA in whole transcriptomic sequencing data | TTMV fusion to a 27 bp exonised RARA intron 2 and RARA exon 3 | NA | NA | NA | 549 amino acids |
| Chen et al. 2023.[15] | Cluster analysis followed by manual investigation of RARA in whole transcriptomic sequencing data and whole genome sequencing data | NA | 2110 bp insertion | NA | NA | 485 amino acids |

NA: data not available

### Supplementary Figure 1

A. SRR14060870

(RNA-based *RARA*::TTMV::*RARA* contig#2 from Trinity; length=752 bp)

CGGGTGGGCTGACCACCCAAACCCCATCTGGGCCCAGGCCCC**C**TGCCCCGAGGAGGGGTGGTCTGAAGCCCACCAGA
GCCCCCTGCCAGACTGTCTGCCTCCCTTCTGACTGTGGCCGCTTGGCATGGCCAGCAACAGCAGCTCCTGCCCGACA
CCTGGGGGCGGGCACCTCAATGGGTACCCGGTGCCTCCCTACGCCTTCTTCTTCCCCCCTATGCTGGGTGGACTCTC
CCCGCCAGGCGCTCTGACCACTCTCCAGCACCAGCTTCCAGTTAGTGGATATAGCACACCATCCCCAGCCAATGTCA
AGACTTCAACCTGTAAAAACTTC**C**AAAAGAAACCAACGCTTAGACTGGATTAATGGCATCGTCCAGATACACAACTT
AATCTGCGGCTGTGAAAAACCTCTAAAACACACCATTGAAGAAATTTGGGCTCA**G**GAACCAAGCCTACATCCCTATC
ACCAATCATGCCCTGGTACTGGAA**G**CG**A**AGACCATACTGGA**C**ACGTCGTCCTTCTCCCCAAATGTGGGTGGGGTGCC
CACATTTCAGCCATTGAGACCCAGAGCAGCAGTTCTGAAGAGATAGTGCCCAGCCCTCCCTCGCCACCCCCTCTACC
CCGCATCTACAAGCCTTGCTTTGTCTGTCAGGACAAGTCCTCAGGCTACCACTATGGGGTCAGCGCCTGTGAGGGCT
GCAAGGGCTTCTTCCGCCGCAGCATCCAGAAGAACATGGTGTACACGTGTCACCGGGAC

*RARA* exon 2 - chr17:38487347-38487648 (301/302) [99.7%]

TTMV MN771920.1 - 492-699 (203/208) [98%]

*RARA* intron 2 - chr17:38490465-38490502 (38/38) [100%]

*RARA* exon 3 - chr17:38504568-38504716 (149/149) [100%]

*RARA* exon 4 - chr17:38506036-38506089 (54/54) [100%]

(RNA-based *RARA*::TTMV::*RARA* contig#10 from Trinity; length=816 bp)

CGGGTGGGCTGACCACCCAAACCCCATCTGGGCCCAGGCCCC**C**TGCCCCGAGGAGGGGTGGTCTGAAGCCCACCAGA
GCCCCCTGCCAGACTGTCTGCCTCCCTTCTGACTGTGGCCGCTTGGCATGGCCAGCAACAGCAGCTCCTGCCCGACA
CCTGGGGGCGGGCACCTCAATGGGTACCCGGTGCCTCCCTACGCCTTCTTCTTCCCCCCTATGCTGGGTGGACTCTC
CCCGCCAGGCGCTCTGACCACTCTCCAGCACCAGCTTCCAGTTAGTGGATATAGCACACCATCCCCAGCCATTTATG
CCGCCAGACGGAGACGCGAAAGCAACTTTCAGCGGCTTAGCCTGGGCGGGTGCCGAAGATGTCAAGACTTCAACCTG
TAAAAACTTC**C**AAAAGAAACCAACGCTTAGACTGGATTAATGGCATCGTCCAGATACACAACTTAATCTGCGGCTGT
GAAAAACCTCTAAAACACACCATTGAAGAAATTTGGGCTCA**G**GAACCAAGCCTACATCCCTATCACCAATCATGCCC
TGGTACTGGAA**G**CG**A**AGACCATACTGGA**C**ACGTCGTCCTTCTCCCCAAATGTGGGTGGGGTGCCCACATTTCAGCCA
TTGAGACCCAGAGCAGCAGTTCTGAAGAGATAGTGCCCAGCCCTCCCTCGCCACCCCCTCTACCCCGCATCTACAAG
CCTTGCTTTGTCTGTCAGGACAAGTCCTCAGGCTACCACTATGGGGTCAGCGCCTGTGAGGGCTGCAAGGGCTTCTT
CCGCCGCAGCATCCAGAAGAACATGGTGTACACGTGTCACCGGGAC

*RARA* exon 2 - chr17: 38487347-38487648 (301/302) [99.7%]

TTMV MN771920.1 – 340-400 (60/61) [98%] + 490-699 (205/210) [98%]#

*RARA* intron 2 - chr17:38490465-38490502 (38/38) [100%]

*RARA* exon 3 - chr17:38504568-38504716 (149/149) [100%]

*RARA* exon 4 - chr17:38506036-38506089 (54/54) [100%]

#See Supplementary Figure 2A for manual merging of blastn alignments

(RNA-based *RARA*::TTMV contig#13 from Trinity; length=250 bp)

CAGACACTCTGTCCATCTGGAGCCTGGGCTCATTGGAGGGTTGGAGGTGGGGGCTGGGGCCTGGCCCCATCCTTGCC
CATCCTGGCCAGAGGGAGCAAATGCTTTGCCTTCCCCTCCCCTCAATGTTGCAAGCTTCTTCCTCTTTCTTAGAGGT
GGGTGGGGAA**G**GTTCAGGAACCGAATCCTCAGACTATTAACAGAGGAAAGTTCATAAACCTTGGCTGTGTACAAACT
GCTCTTTAGGAAATAAAAG

*RARA* intron 2 - chr17:38490283-38490448* (166/166) [100%]

TTMV MN771920.1 – 2629-2704* (75/76) [99%]

*includes shared AAGG (4 bp)

B. SRR14060871

(RNA-based *RARA*::TTMV::*RARA* contig#1 from Trinity; length=977 bp)

```
CCCATGCCCCGAGGAGGGGTGGTCTGAAGCCCACCAGAGCCCCCTGCCAGACTGTCTGCCTCCCTTCTGACTGTGGC
CGCTTGGCATGGCCAGCAACAGCAGCTCCTGCCCGACACCTGGGGGCGGCACCTCAATGGGTACCCGGTGCCTCCC
TACGCCTTCTTCTTCCCCCCTATGCTGGGTGGACTCTCCCCGCCAGGCGCTCTGACCACTCTCCAGCACCAGCTTCC
AGTTAGTGGATATAGCACACCATCCCCAGCCAATGTCAAGACTTCAACCTGTAAAAACTTCTAAAAGAAACCAACGC
TTAGACTGGATTAATGGCATCGTCCAGATACACAACTTAATCTGCGGCTGTGAAGAACCTCTAAAACACACCATTGA
AGAAATTTGGGCTCAAGAACCAAGCCTACATCCCTATCACCAATCATGCCCTGGTACTGGAAACGGAGACCATACTG
GAGACGTCGCAGAACTAGAAAATGGGGATTTAGACCGTTTGTTCGCCGACGACTTTGGAGAAGAAGACGCAGGCACC
AGTACAGGGAGCAGACCCCTTTCTCCCAACACCCCAAGAAGCAGCACCGACACAAGGCGTTCTACTGCCTCAGCCTC
TCGAGTAGCTGGGACTACAGCCATTGAGACCCAGAGCAGCAGTTCTGAAGAGATAGTGCCCAGCCCTCCCTCGCCAC
CCCCTCTACCCCGCATCTACAAGCCTTGCTTTGTCTGTCAGGACAAGTCCTCAGGCTACCACTATGGGGTCAGCGCC
TGTGAGGGCTGCAAGGGCTTCTTCCGCCGCAGCATCCAGAAGAACATGGTGTACACGTGTCACCGGGACAAGAACTG
CATCATCAACAAGGTGACCCGGAACCGCTGCCAGTACTGCCGGCTGCAGAAGTGCTTTGAAGTGGGCATGTCCAAGA
AGTCTGTGAGAAACGACCGAAACAAGAAGAAGAAGGAGGTGCCCACGCCCGAG
```

*RARA* exon 2 - chr17:38487386-38487648 (263/263) [100%]

TTMV MN769771.1 – 438-722* (285/285) [100%] + 2456-2502*^ (47/47) [100%]#

*RARA* intron 2 - chr17:38489987-38490031^ (45/45) [100%]

*RARA* exon 3 - chr17:38504568-38504716 (149/149) [100%]

*RARA* exon 4 - chr17:38506036-38506177 (140/142) [99%]

*RARA* exon 5 - chr17:38508162-38508211 (49/50) [98%]

*includes shared GG (2 bp)

^includes shared CA (2 bp)

#See Supplementary Figure 2B for manual merging of blastn alignments

**Supplementary Figure 1.** Fusion contigs derived from public RNA-seq data of the inaugural TTMV::RARA publication. (A) SRR14060870: Trinity generated several fusion contigs, where the predominant (top) was a RARA::TTMV::RARA contig (length 752 bp) characterized by an aberrant 274 bp insertion (209 bp with high sequence identity to the TTMV isolate MN771920.1 and 38 bp from RARA intron 2) between RARA exons 2 and 3, thus disrupting the standard RARA frame and predicted to instead utilize an ATG start codon of TTMV sharing the frame of RARA exon 3 (see **Supplementary Figure 2A**). The TTMV to RARA intron 2 connection was inferred to represent the underlying DNA breakpoints from the 3' side of TTMV viral integration. A minor fusion contig (middle) had a slightly longer TTMV insertion with an additional 62 bp at the 5' end, most consistent with alternative splicing. Another minor fusion contig (bottom) was predicted to represent the underlying DNA breakpoints from the 5' side of TTMV viral integration. (B) SRR14060871: Trinity generated a RARA::TTMV::RARA contig of length 977 bp, characterized by an aberrant 373 bp insertion (328 bp with high sequence identity to 2 regions of the TTMV isolate MN769771.1 and 45 bp from RARA intron 2) between RARA exons 2 and 3, again disrupting the standard RARA frame and predicted to instead utilize an ATG start codon within TTMV sharing the frame of RARA exon 3 (see **Supplementary Figure 2B**). The TTMV to RARA intron 2 connection was again inferred to represent the underlying DNA breakpoints from the 3' side of TTMV viral integration. Split-reads were predicted to capture the underlying DNA breakpoints from the 5' side of TTMV viral integration (see **Supplementary Table 2**) but were not part of a Trinity contig.

**Supplementary Figure 2**

A. SRR14060870 (blastn and blastx alignments [MN771920.1])

```
                M  S  R  L  Q  P  V  K  T  S  K  R  N  Q  R  L  D  W  I  N  G  I
          ATGTCAAGACTTCAACCTGTAAAAACTTCCAAAAGAAACCAACGCTTAGACTGGATTAATGGCATCG
...ATTTTAGATGTCAAGACTTCAACCTGTAAAAACTTCTAAAAGAAACCAACGCTTAGACTGGATTAATGGCATCG

V  Q  I  H  N  L  I  C  G  C  E  K  P  L  K  H  T  I  E  E  I  W  A  Q  E  P
TCCAGATACACAACTTAATCTGCGGCTGTGAAAAACCTCTAAAACACACCATTGAAGAAATTTGGGCTCAGGAACCA
TCCAGATACACAACTTAATCTGCGGCTGTGAAAAACCTCTAAAACACACCATTGAAGAAATTTGGGCTCAAGAACCA

 S  L  H  P  Y  H  Q  S  C  P  G  T  G  S  E  D  H  T  G  H  V
AGCCTACATCCCTATCACCAATCATGCCCTGGTACTGGAAGCGAAGACCATACTGGACACGTCG
AGCCTACATCCCTATCACCAATCATGCCCTGGTACTGGAAACGCAGACCATACTGGAGACGTCGCAGAACTAGA...
```

```
Blastx
SRR14060870      5 QPVKTSKRNQRLDWINGIVQIHNLICGCEKPLKHTIEEIWAQEPSLhPYHQSC
Cdd:pfam02957    2 RPPRYNPKGLELQWINSIVRSHDLFCGCGDPLKHLIHPILRQLPAL-PAAPEE

SRR14060870        PG--------TGSEDHTG 67
Cdd:pfam02957      PGdlakwlttTGEDGGTG 71
```

B. SRR14060871 (blastn merging and blastx alignments [MN769771.1])

```
             M  S  R  L  Q  P  V  K  T  S  K  R  N  Q  R  L  D  W  I  N  G  I  V
       ATGTCAAGACTTCAACCTGTAAAAACTTCTAAAAGAAACCAACGCTTAGACTGGATTAATGGCATCGTC
...TTTAGATGTCAAGACTTCAACCTGTAAAAACTTCTAAAAGAAACCAACGCTTAGACTGGATTAATGGCATCGTC

 Q  I  H  N  L  I  C  G  C  E  E  P  L  K  H  T  I  E  E  I  W  A  Q  E  P  S
CAGATACACAACTTAATCTGCGGCTGTGAAGAACCTCTAAAACACACCATTGAAGAAATTTGGGCTCAAGAACCAAG
CAGATACACAACTTAATCTGCGGCTGTGAAGAACCTCTAAAACACACCATTGAAGAAATTTGGGCTCAAGAACCAAG

  L  H  P  Y  H  Q  S  C  P  G  T  G  N  G  D  H  T  G  D  V  A  E  L  E  N
CCTACATCCCTATCACCAATCATGCCCTGGTACTGGAAACGGAGACCATACTGGAGACGTCGCAGAACTAGAAAATG
CCTACATCCCTATCACCAATCATGCCCTGGTACTGGAAACGGAGACCATACTGGAGACGTCGCAGAACTAGAAAATG

G  D  L  D  R  L  F  A  D  D  F  G  E  E  D  A  G  T  S  T  G
GGGATTTAGACCGTTTGTTCGCCGACGACTTTGGAGAAGAAGACGCAGGCACCAGTACAGG---------- .....
GGGATTTAGACCGTTTGTTCGCCGACGACTTTGGAGAAGAAGACGCAGGCACCAGTACAGGGTAAGAAGAC .....

                          S  R  P  L  S  P  N  T  P  R  S  S  T  D  T
.............  ----------GAGCAGACCCCTTTCTCCCAACACCCCAAGAAGCAGCACCGACACA
(~1.7kb).....  GAATTGCAGGAGCAGACCCCTTTCTCCCAACACCCCAAGAAGCAGCACCGACACAGGACT...
```

```
Blastx
SRR14060871      5 QPVKTSKRNQRLDWINGIVQIHNLICGCEEPLKHTIEEIWAQEPSLhPYHQSC
Cdd:pfam02957    2 RPPRYNPKGLELQWINSIVRSHDLFCGCGDPLKHLIHPILRQLPAL-PAAPEE

SRR14060871        PG--------TG--------NGDHT-GDVA--ELENGDLDRLFADDFGEE 88
Cdd:pfam02957      PGdlakwlttTGedggtgprGGDGTaGDAAgeGLDEGDLDLLFAEDFAED 103
```

C. SRR14060870 (Minor contig: merging of blastn alignments [MN771920.1])

```
        TTTATGCCGCCAGACGGAGACGCGAAAGCAACTTTCAGCGGCTTAGCCTGGGCGGGTGCCGAAG-----
...CTGAGTTTATGCCGCCAGACGGAGACGCGAAAGGAACTTTCAGCGGCTTAGCCTGGGCGGGTGCCGGAGGTGAG

-----------------------------------------------------------------------------
TTTACCACCGTAGTCAAGGGGCAATTCGGGCTGGCTAAGTCTGGCGGAACGGGCAAGAAACTTAAAAAATATTTTTA

------ATGTCAAGACTTCAACCTGTAAAAACTTCCAAAAGAAACCAACGCTTAGACTGGATTAATGGCATCGTCCA
TTTTAGATGTCAAGACTTCAACCTGTAAAAACTTCTAAAAGAAACCAACGCTTAGACTGGATTAATGGCATCGTCCA

GATACACAACTTAATCTGCGGCTGTGAAAAACCTCTAAAACACACCATTGAAGAAATTTGGGCTCAGGAACCAAGCC
GATACACAACTTAATCTGCGGCTGTGAAAAACCTCTAAAACACACCATTGAAGAAATTTGGGCTCAAGAACCAAGCC

TACATCCCTATCACCAATCATGCCCTGGTACTGGAAGCGAAGACCATACTGGACACGTCG
TACATCCCTATCACCAATCATGCCCTGGTACTGGAAACGCAGACCATACTGGAGACGTCGCAGAACTAGA...
```

**Supplementary Figure 2.** TTMV components of the fusion contigs from the inaugural TTMV::*RARA* publication. (A-B) In both SRR14060870 (A) and SRR14060871 (B), *RARA* exon 2 was directly spliced to an open reading frame of TTMV containing a putative conserved domain from the TT viral ORF2 superfamily by blastx and sharing the same frame as *RARA* exon 3. In SRR14060871 (B), there was also inferred splicing of ~1.7 kb from TTMV, thereby joining TTMV ORF2 to a short 46 bp out-of-frame segment within TTMV ORF1 followed by fusion to a short 45 bp segment of *RARA* intron 2 and cryptic splicing to *RARA* exon 3 (see **Supplementary Figure 1B**). Thus, SRR14060871 was an example where the DNA breakpoint of the 3' side of the TTMV integration was predicted to occur within ORF1, yet ORF2 was still utilized on the RNA side. (C) Manual merging of the blastn alignments from the minor fusion contig of SRR14060870 was consistent with alternative splicing of RARA exon 2 to an earlier putative splice acceptor site within TTMV as well as splicing of an 88 bp segment from TTMV directly before the ATG start codon of the ORF containing the putative conserved domain from the TT viral ORF2 superfamily, overall similar to the findings of cases 1-2 (see **Figure 2C-D**).

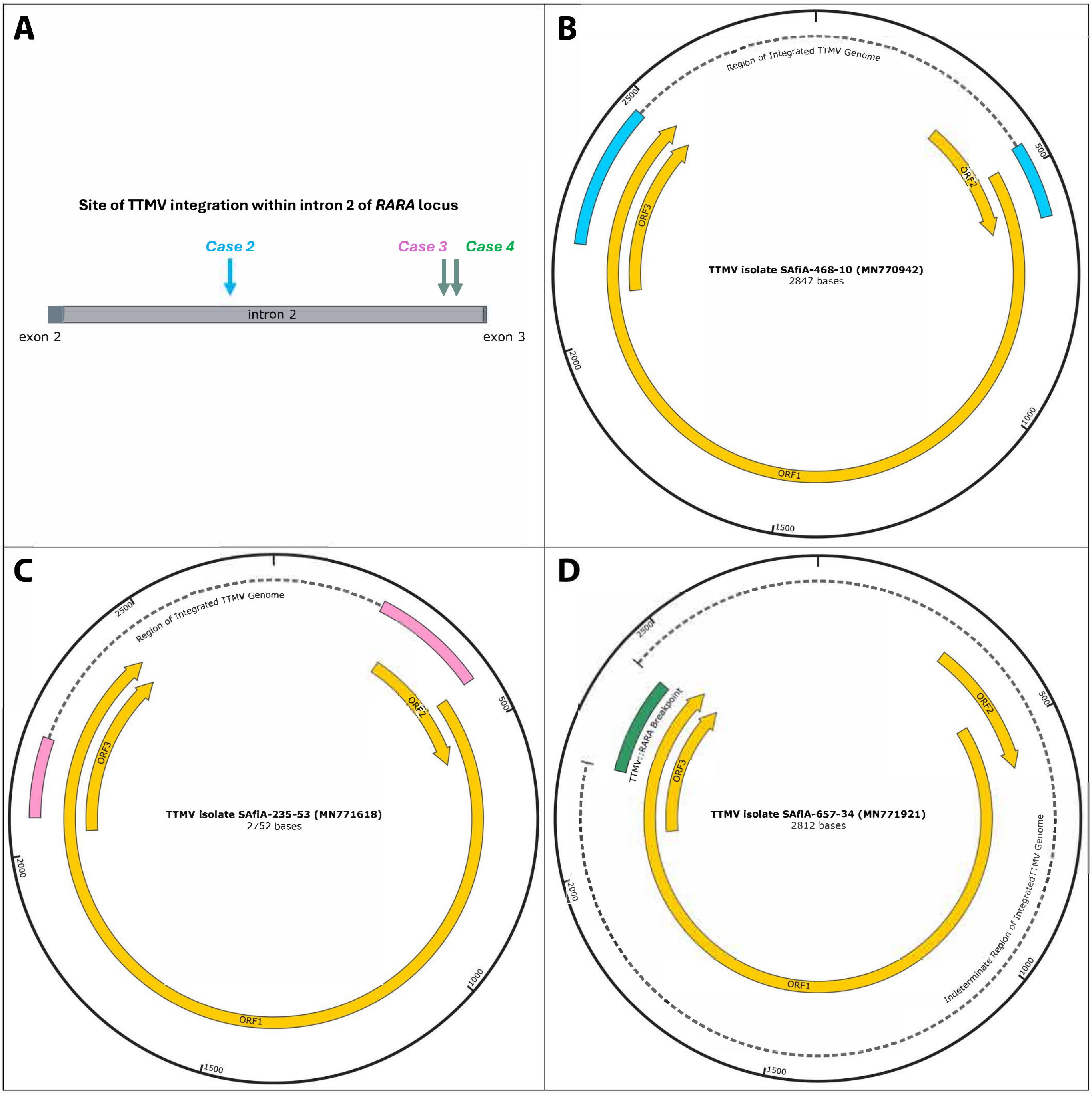


**Supplementary Figure 3:** Recurrent partial integration of TTMV within intron 2 of RARA. A) Schematic of breakpoint cluster region (intron 2, light grey solid bar) of the RARA gene. Exons 2 and 3 shown as dark gray solid bars. Three arrows indicate the sites of partial TTMV integration identified for cases 2-4; arrow color denotes case and corresponding TTMV isolate genome with contig blastn alignments (cyan, case 2, panel B; pink, case 3, panel C; green, case 4, panel D). B-D) Visualization of the labeled TTMV isolate genome with annotated encoded proteins ORFl-3 in gold. The colored solid bars indicate the portion of the TTMV genome that aligns via blastn with the assembled contigs for cases 2 -4. For cases 2 and 3 (panels Band C, respectively), these correspond to RARA::TTMV breakpoint (left solid bar, end of ORF I/overlapping ORF3) and TTMV::RARA breakpoint (right solid bar, ORF2), whereas, in case 4 (panel D), only the TTMV::RARA breakpoint was identified (i.e. single green solid bar, end of ORF I/overlapping ORF3). For panels Band C, the dotted line between the colored solid bars represents the remainder of the TTMV genome that is inferred to have been linearized and integrated within RARA intron 2. As only one breakpoint was identified for case 4, the dotted line in panel D represents an indeterminate remainder of the viral genome that could be present in the patient's genome. Images were generated using SnapGene® software from Dotmatics (available at snapgene.com).

**Supplementary Figure 4**

A. Case 3 (blastx of TTMV::*RARA* contig from DNA)

```
Case3_DNA       22 PIKTSARNRRLDWINGIVHLHNLVCGCDKPLEHTLEEIYTQEPQL 66
Cdd:pfam02957    3 PPRYNPKGLELQWINSIVRSHDLFCGCGDPLKHLIHPILRQLPAL 47
```

B. Case 4 (blastx of TTMV::*RARA* contig from DNA)

```
No putative conserved domains detected

Top blastx alignment - MAG: ORF1 [TTV-like mini virus] (68/68) [100%]

Case4_DNA        3 KFLYSFDWRRGYLTQKATTRISADESTEKNVFGPTGFNLFNPQPTQDsse
UGV36713.1     587 KFLYSFDWRRGYLTQKATTRISADESTEKNVFGPTGFNLFNPQPTQDSSE

Case4_DNA          eteteekseketLQILLK  206
UGV36713.1         ETETEEKSEKETLQILLK  654
```

**Supplementary Figure 4.** Blastx of TTMV::RARA contigs from DNA. (A) Case 3: blastx detected the same putative conserved domain from the TT viral ORF2 superfamily as cases 1-2 (B) Case 4: blastx detected no putative conserved domains, however yielded an alignment to TT viral ORF1 (pfam02956) of UGV36713.1 with 100% (68/68) identity on the protein level.
